# Maternal *IGHG* locus duplications impair infants’ passive immunity

**DOI:** 10.1101/2025.10.09.25337669

**Authors:** Ioannis Belios, Dimitra E. Zazara, Ioannis Evangelakos, Julia Hambach, Tom Siegl, Kristoffer Riecken, Marie Albrecht, Christian Müller, Michael Spohn, Agnes Wieczorek, Katrin Rading, Kristin Thiele, Anastasios D. Giannou, Sotirios G. Zarogiannis, Natalie Ledée, Wenjun Wang, Steinar Gijze, Jan Nouta, Karen Manalastas-Cantos, Isabel Graf, Christopher Urbschat, Ann-Christin Tallarek, Maya Topf, Felix Stahl, Friedrich Koch-Nolte, Marc Lütgehetmann, Manfred Wuhrer, Malik Alawi, Martin Becker, Marylyn M. Addo, Anke Diemert, Christian Schlein, Petra C. Arck

**Affiliations:** Division for Experimental Feto-Maternal Medicine, Department of Obstetrics and Fetal Medicine, University Medical Center of Hamburg-Eppendorf (UKE), Hamburg, Germany; Hamburg Center of Translational Immunology, UKE, Hamburg, Germany; Junior Research Center for Reproduction: Sexual and Reproductive Health in Overweight and Obesity, UKE, Hamburg, Germany; University Children’s Hospital, UKE, Hamburg, Germany; Institute of Human Genetics, UKE, Hamburg, Germany; Institute of Immunology, Center for Diagnostics, UKE, Hamburg, Germany; Institute for Visual and Analytic Computing, University of Rostock, Rostock, Germany; Center for Oncology, Department of Stem Cell Transplantation, UKE, Hamburg, Germany; Bioinformatics Core Unit, UKE, Hamburg, Germany; Department of General, Visceral and Thoracic Surgery, UKE, Hamburg, Germany; Section of Molecular Immunology and Gastroenterology, I. Department of Medicine, UKE, Hamburg, Germany; Department of Physiology, Faculty of Medicine, University of Thessaly, Larissa, Greece; Centre d’Assistance Médical á la Procréation Bluets-Drouot, Hôpital Les Bluets, Paris, France; Center for Proteomics and Metabolomics, Leiden University Medical Center, Leiden, The Netherlands; Department of Integrative Virology, Leibniz-Institute for Virology, Hamburg, Germany; Institute for Molecular Virology and Tumor Virology, UKE, Hamburg, Germany; Department of Obstetrics and Fetal Medicine, UKE, Hamburg, Germany; Center for Diagnostics, Institute of Medical Microbiology, Virology and Hygiene, UKE, Hamburg, Germany; Department of Mathematics and Computer Science, Marburg University, Marburg, Germany; Institute for Infection Research and Vaccine Development, Center for Internal Medicine, UKE, Hamburg, Germany; Department for Clinical Immunology of Infectious Diseases, Bernhard Nocht Institute for Tropical Medicine, Hamburg, Germany; Division for Midwifery, Department of Obstetrics and Fetal Medicine, UKE, Hamburg, Germany; German Center for Child and Adolescent Health, Partner Site Hamburg

## Abstract

Infants depend on passive immunity to safeguard them against infections during the first months of life. Maternal immunoglobulin G (IgG) antibodies are actively transported across the placenta and confer this protection. In this study, we discovered common, but previously unrecognized, naturally occurring gene fusions between loci encoding IgG1 and IgG4 subclasses that impair the transplacental IgG transport. These gene fusions result from gene duplications combining regulatory elements of the *Immunoglobulin Heavy Constant Gamma* (*IGHG1)* gene with *IGHG4*-like constant regions. Mothers with these duplications generate antibodies that are less efficiently transferred to the fetus, resulting in lower antibody levels in newborns and a higher risk of respiratory infections during infancy. Our insights warrant consideration in the development of personalized vaccination strategies during pregnancy to better protect infants against infectious diseases.

## Introduction

Early-life infections are a significant cause of death and disease worldwide (1, 2). During this vital period, infants can be protected by passive immunity acquired before birth. Passive immunity is conferred by maternal pathogen-specific immunoglobulin G (IgG) antibodies that cross the placenta to reach the fetus during pregnancy (3). Studies consistently show that higher maternal IgG levels in newborns are linked to fewer infections in infancy (4, 5). The transplacental antibody transfer involves an active transport system, where maternal antibodies bind to a specialized receptor known as the neonatal Fc receptor (FcRn) (*6*). Notably, among the four IgG subclasses, IgG1 is distinguished by its superior transplacental transport efficiency (*7*).

The first recorded evidence of active IgG transfer through the placenta dates to 1966 (*8*). Since then, it has been consistently observed that the transplacental transfer rate of IgGs varies considerably between mother/neonate pairs. (*9*–*15*) (fig. S1). In our prior investigations, we observed that the interindividual variability in IgG transfer efficiency is irrespective of the pathogen-specificity of the antibody (*16*). To assess whether maternal or placental factors determine IgG transfer efficiency between mother/neonate pairs, we monitored the transplacental transfer rate across multiple pregnancies in the same women. This approach facilitated the identification of common structural variants within the *IGHG* locus, which generate chimeric *IGHG1/IGHG4* alleles. *IGHG 1* encodes the constant region of the IgG1 heavy chain, which is involved in antibody functions like complement activation and interaction with immune cells. The variants we identified account for a substantial proportion of the interindividual variability of the transplacental IgG transfer, offering valuable insights into the mechanisms by which some mothers consistently confer higher or lower levels of antibody-mediated protection to their offspring.

## Results

### Maternal factors determine transplacental IgG transfer efficiency across pregnancies

To investigate the factors influencing the interindividual variability in transplacental IgG transfer, we analyzed mothers participating in the population-based, prospective, longitudinal pregnancy study ‘PRINCE’ (Prenatal Identification of Children’s Health) with two consecutive pregnancies (Fig. 1A). We quantified the transplacental antibody transfer rate (TPTR) for eight pathogen-specific IgG antibodies by examining maternal peripheral venous serum and neonatal cord blood samples (Fig. 1B). All eight pathogens selected are vaccine preventable. The pathogen-specific TPTRs were normalized into z-scores corresponding to each mother-neonate pair during each pregnancy (Fig. 1C). Clustering analysis revealed that mother/neonate pairs exhibiting high TPTR z-scores in the first pregnancy maintained similarly elevated scores in the subsequent pregnancy (Fig. 1C). In contrast, pairs with low initial scores sustained low scores in the subsequent pregnancy (Fig. 1C). Additionally, a subset of mother-neonate pairs displayed intermediate TPTR z-score values across both pregnancies. One woman received a booster vaccination against B. pertussis and Tetanus, which was easily detectable during the second pregnancy.

**Fig. 1.**
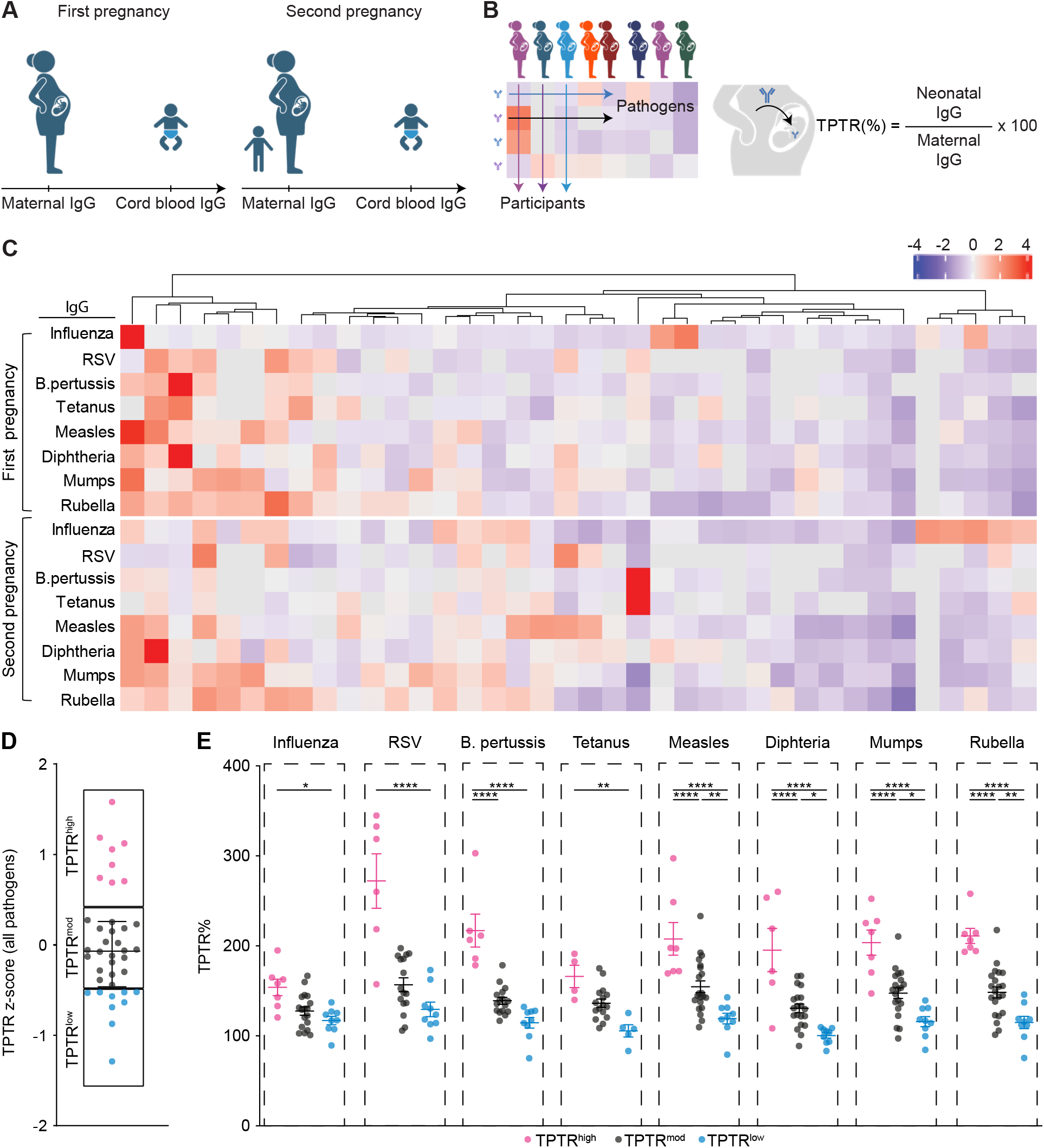
Efficacy of transplacental immunoglobulin G (IgG) transfer across successive pregnancies in the same mother. (**A**) Schematic representation delineating the study design. (**B**) Illustration of the adopted methodology used to depict the TPTR of pathogen-specific IgG, along with the corresponding formula for its calculation. The color scale represents the TPTR z-score, indicating the deviation from the mean for each row. In the miniature heat map, each column represents the TPTR for all pathogen-specific IgG measured within a single mother/infant (neonate) pair. Each row depicts the TPTR for a specific pathogen across all participating mother/infant pairs. (**C**) Heat maps illustrating the z-scores of the pathogen-specific IgG TPTR for each mother/infant pair during both the first and second pregnancies. The data is presented in accordance with the methodology outlined in (B). (**D**) Dot plot illustrating the interquartile categorization of mother/infant pairs into three categories: lowest (TPTR^low^), moderate (TPTR^mod^), and highest (TPTR^high^), derived from the combined TPTR z-scores across all pathogen-specific antibodies assessed during first and second pregnancies. (**E**) Absolute combined TPTR values across quartiles of mother-infant pairs. For each individual mother-infant pair, the combined mean TPTR of the first and second pregnancy was computed. Data are expressed as mean ± standard error of the mean (SEM). *;p < 0.05, **; p < 0.01; ***;p<0.001, ****;p<0.0001, as assessed by One-way ANOVA with multiple comparisons.

To validate our findings, we calculated composite TPTR z-scores encompassing all antibodies and pregnancies (TPTR z-score (all pathogens)). The cohort was stratified into quartiles, merging the two middle quartiles. Mother/neonate pairs in the lowest 25% were classified as TPTR^low^ quartile (n=9), those in the highest quartile as TPTR^high^ (n=8) and the remaining two middle quartiles as TPTR^mod^ (n=21) (Fig. 1D). We subsequently illustrated the combined mean TPTR from the first and second pregnancies for every individual for each pathogen-specific antibody, confirming that mother/neonate pairs in the TPTR^high^ group exhibited significantly elevated TPTR in both pregnancies, whilst pairs in the TPTR^low^ quartile presented lower TPTR in both pregnancies (Fig. 1E). No significant differences in maternal demographics or neonatal anthropometric parameters were observed between the two pregnancies, except the expected increase in maternal age and body mass index (table S1). Seropositivity data for maternal and cord blood samples are provided in table S2, with antibody titers and maternal-cord blood correlations illustrated in figs. S2, S3, and S4. Collectively, these results indicate that interindividual differences in transplacental IgG transfer are independent of placental or neonatal parameters, but instead are predominantly influenced by stable maternal factors. Overall, these findings suggest that the interindividual variability in IgG transfer efficiency is not driven by placental or neonatal factors but mainly determined by stable maternal characteristics.

### Structural IGHG variants create chimeric IGHG1/IGHG4 alleles associated with reduced TPTR

Based on this observation, we hypothesized that maternal genetic factors determine the transplacental IgG transfer efficiency. Previous research unrelated to pregnancy shows that specific IgG1 allotypes bind differentially to the FcRn receptor *in vitro* (*17*), prompting us to analyze *IGHG1* gene variants in our study (Fig. 2A). Strikingly, Sanger sequencing using established primers for *IGHG1* locus (*18*) revealed *IGHG4*-like sequence signatures within the *IGHG1*-targeted amplicons in several cases (exemplarily shown in Fig. 2B, Sanger sequencing results fig. S5A, table S3).

**Fig. 2.**
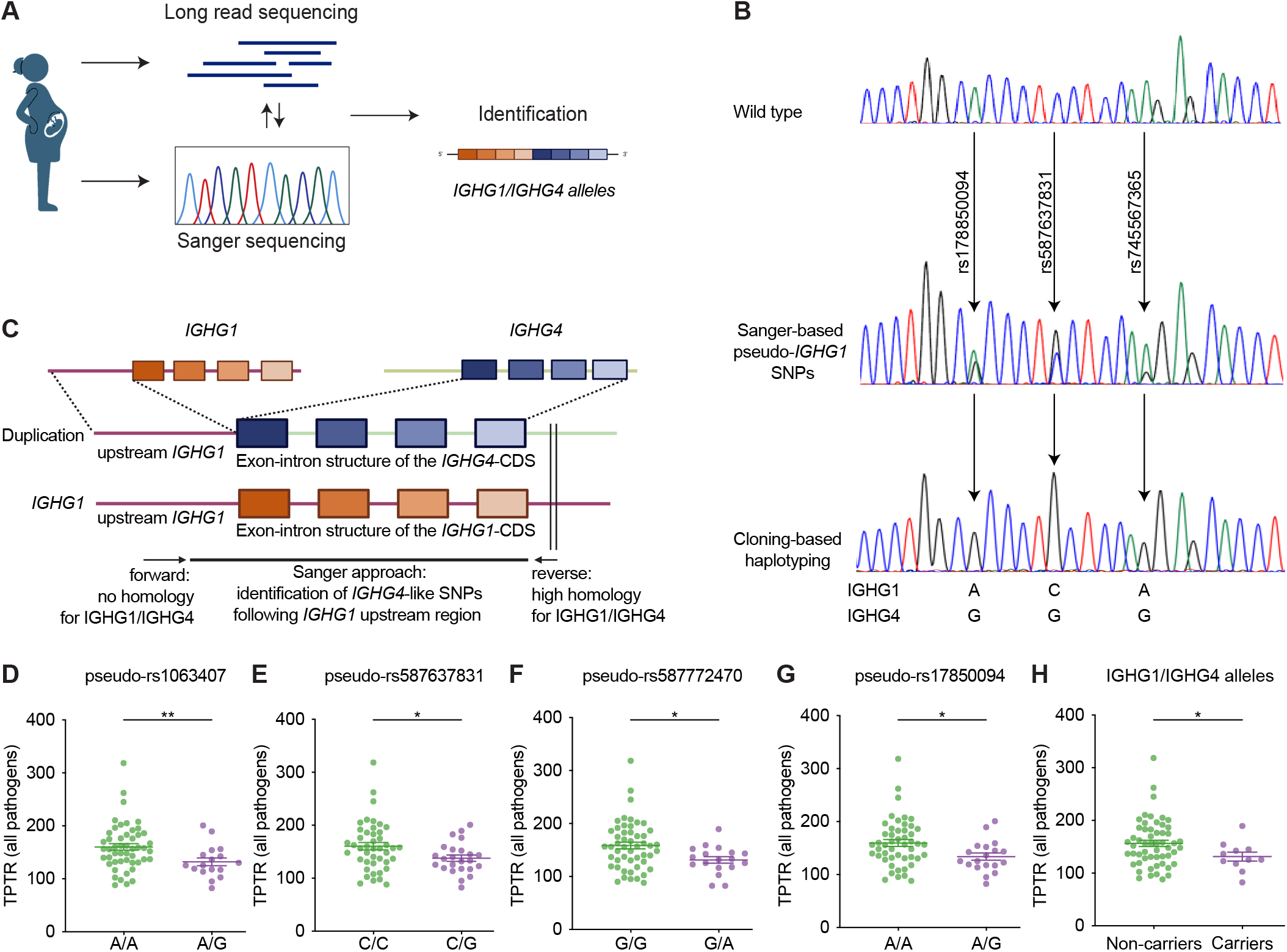
Structural IGHG variants create chimeric IGHG1/IGHG4 alleles associated with reduced TPTR. **(A**) Schematic overview of the sequencing strategy and analysis pipeline used to identify structural variation within the IGHG locus in the study cohort. (**B**) Representative Sanger sequencing traces and clone-based haplotyping illustrating the detection of IGHG1/IGHG4 chimeric alleles (see also fig. S6). Sanger sequencing data were compared with the reference sequences of *IGHG1* and *IGHG4*. (**C**) Proposed gene architecture of duplications, resulting in IGHG1/IGHG4 alleles. (**D-G**) Comparison of the TPTR across all pathogens between individuals carrying at least one alternate allele at each of the four pseudo-SNP markers most strongly associated with reduced transfer efficiency. Data were analyzed using the Welch’s test. *; p value<0.05, **; p < 0.01. Participants characterized as wild type are depicted in green, individuals with a mixed *IGHG4*-like signal in Sanger sequencing are marked in purpple. **(H)** TPTR comparison in mothers carrying all four pseudo-SNP markers simultaneously versus non-carriers. *; p value<0.05, as assessed by Welch’s test

Haplotyping through PCR amplicon cloning confirmed these as distinct haplotypes (Fig. 2B). To distinguish between nonspecific amplification of canonical *IGHG4* and *IGHG1* loci harboring *IGHG4*-like variants, we performed High Fidelity long-read sequencing in a subgroup of women with Sanger-confirmed complex haplotypes. Since alignment to GRCh38 (the reference human genome assembly maintained by the Genome Reference Consortium) did not allow to recover reads bearing the suspected SNP patterns, we conducted *de novo* assembly followed by *in silico* PCR using the same primers. Intriguingly, these assemblies revealed duplications and triplications of the *IGHG* region with a hybrid genomic structure. Specifically, we detected an upstream region resembling *IGHG1*, which is fused to constant regions exhibiting *IGHG4*-like exon-intron architecture (Fig. 2C, fig. S6A, B, C, table S4).

Recent studies identified IGHG locus duplications as *IGHG4* expansions with high allele frequency in European populations (*19, 20*). However, our data show that these alleles also include *IGHG1*-derived upstream elements (Fig. 2C, fig. S6A), making them functionally distinct. The 5′ end upstream sequence plays a key role in regulating expression and isotype switching. Thus, we propose that the structural hybrid variants we identified produce chimeric IgG1/IgG4 antibodies with modified functions. Although the canonical *IGHG1* copy sustained a complete homology among the individuals (table S5), detailed analysis of the duplications revealed a slight heterogeneity within the detected duplications, especially in the hinge exon (table S6). Importantly, in a triplication found in one woman, the hinge exon itself displayed an intra-exonic hybrid *IGHG1/4* structure (Fig. S6C).

We observed distinct variants in Sanger sequencing traces, which allowed us to adapt this method for cost-efficient detection of TPTR-associated marker SNPs. All observed variants in the cohort were annotated relative to the *IGHG1* reference sequence, identifying 32 pseudo-SNPs (19 missense) (fig. S5A, table S3). However, not all SNPs were similarly detected in all individuals with a putative duplication, supporting heterogeneity within the duplications. Therefore, we conducted a regression analysis with the assumption of an additive genetic model of the presence of a specific pseudo-SNP to the all-pathogen TPTR to define a subset of candidate marker SNPs predictive of low TPTR. This revealed four significant pseudo-SNPs (pseudo-rs17850094, pseudo-rs587637831, pseudo-rs587772470, pseudo-rs1063407) (table S3). The TPTR across all pathogens was also significantly lower in the individuals carrying one of the four identified pseudo-SNPs (Fig. 2D-G). We observed that pregnant non-carriers still show a slightly higher frequency of certain pseudo-SNPs compared to data derived from Genome Aggregation Database (gnomAd, version 4.0.1) (fig. S5B). This may result from the challenges in sequencing this locus in existing datasets or from the high specificity but lower sensitivity of this screening method. Yet, to reduce the risk of false-positive duplication prediction in individuals based on canonical *IGHG1* germline variants, we used the parallel presence of all four “marker pseudo-SNPs”. By this approach, we identified n=11 individuals bearing the four marker pseudo-SNPs with a high likelihood of being carriers for the chimeric *IGHG1/IGHG4* duplications. Strikingly, these pregnant carrier women exhibited significantly reduced TPTR compared to duplication-negative (by the absence of the marker pseudo-SNPs) non-carrier women (n=55) (Fig. 2H).

Importantly, the absolute pathogen-specific IgG titers did not differ between carriers of IGHG locus duplications and non-carriers (fig. S7), nor did their demographic characteristics (table S7). Together, these findings demonstrate that Sanger-based detection of chimeric *IGHG4/IGHG1* duplications offers a practical and cost-efficient proxy for identifying individuals at risk of diminished transplacental IgG transfer.

### Functional analysis of chimeric IgG1/IgG4 antibodies

To assess the potential structural effects of the identified missense pseudo-SNPs within the duplication that differentiates *IGHG1* from *IGHG4*, we mapped these variants onto X-ray structures of the human heavy chain Fc fragment (PDB ID 4N0U) (*21*) and the Fab constant fragment of the human anti-influenza IgG1 (PDB ID 5UGY)(*22*) (Fig. 3A, fig. S8). Of these, 61% of the variants are localized in the Fc region, including one of the marker pseudo-SNPs (rs1063407). The remaining missense pseudo-SNPs that distinguish these sequences from canonical IgG1 are clustered within the strands of the CH1 domain (Fig. 3 C, fig. S8), including the IgG1/IgG4 chimeric marker pseudo-SNPs (rs587637831) (Fig. 3C, fig. S8). None of the variants overlap with the FcRn binding interface, indicating that receptor interactions remain unaffected (*23*) (Fig. 3B-D, S8). However, the overall antibody structure, stability, and functionality could be affected by the chimeric antibody resulting from the presence of the rs1063407 SNP at the inter-heavy chain interactions site of the Fc region (Fig. 3E, F, fig. S8) (*24, 25*).

**Fig. 3.**
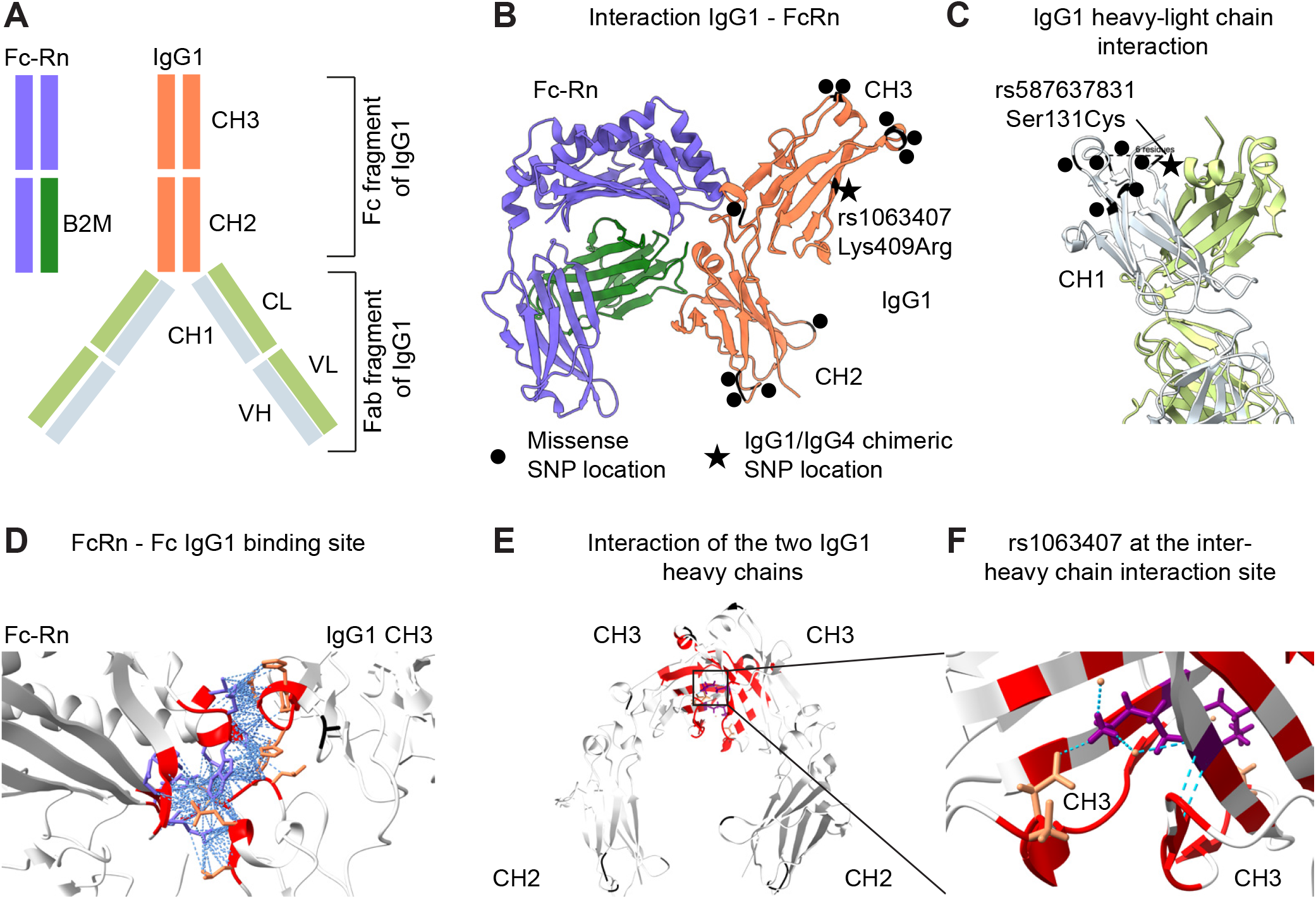
Graphical depiction of IgG three-dimensional protein structures. (**A**) Schematic depiction of Fc-Rn and IgG1 structures. The chosen colors also apply to the structures shown in B and C. (**B**) Ribbon representation of the interaction site between the IgG1 heavy constant chain (marked in orange) and the Fc-Rn (marked in purple; PDB ID: 4N0U). (**C**) Protein structures of the interaction site between the anti-influenza IgG1 heavy chain and the IgG1 light chain (marked in green, PDB ID 5UGY). For (B) and (C), the amino acid positions affected by the specific pseudo-SNPs are highlighted by black dots. The black asterisks highlight the domains of the two missense marker pseudo-SNPs. (**D**) PICKLUSTER, a protein-interface clustering and analysis plug-in for UCSF ChimeraX, version 1.9 (*40,41*) was used to generate a detailed view of the binding site (shown in red) between the IgG1 heavy chain and the Fc-Rn on PDB ID: 4N0U. None of the amino acids affected by SNPs were identified at this site. (**E**-**F**) Using PICKLUSTER on PDB ID:5JII to assess the interaction between two heavy constant chains of IgG1 (shown in red), revealed that the amino acid altered by the missense SNP rs1063407 is involved in the interaction between the two heavy chains (shown in purple), hereby possibly affecting IgG1 stability. All other SNPs were absent from the interaction site. Abbreviations used in (A)– (F): Constant Heavy (CH) 1; the first constant domain of the IgG1 heavy chain, CH2; the second constant domain of the IgG1 heavy chain, CH3; the third constant domain of the IgG1 heavy chain, Variable Heavy (VH); the variable domain of the IgG1 heavy chain, Constant Light (CL); the constant domain of the IgG1 light chain, Variable Light (VL); the variable domain of the IgG1 light chain, Fragment antigen-binding (Fab), Fragment crystallizable (Fc region), B2M; b2-microglobulin.

Another factor influencing the transplacental transfer efficiency could be the Fc glycosylation patterns of maternal IgG (*26*). Therefore, we investigated whether the chimeric antibodies we identified show different Fc glycosylation profiles, using a well-established method (*27*). None of the glycosylation types tested, including fucosylation or bisection, demonstrated any correlation with TPTR across all pathogens (Fig. 4A-C), and glycosylation patterns showed no differences between individuals with or without the IgG1/IgG4 chimeric antibodies (figs. S9, S10). Analyzing the relative abundance of specific glycans on IgG1 in relation to the TPTR revealed that the H4N4F1S1 glycan is negatively correlated with the TPTR of anti-measles, anti-mumps, and anti-tetanus IgG antibodies (Fig. 4D). However, because no significant differences were observed for this glycan between non-carriers and carriers of the chimeric IgG1/IgG4 (Fig. 4E, figs. S9, S10), we conclude that maternal IgG Fc glycosylation patterns do not result or account for the differences in antibody transfer efficiency observed in carriers for the chimeric *IGHG1/IGHG4* duplications.

**Fig. 4.**
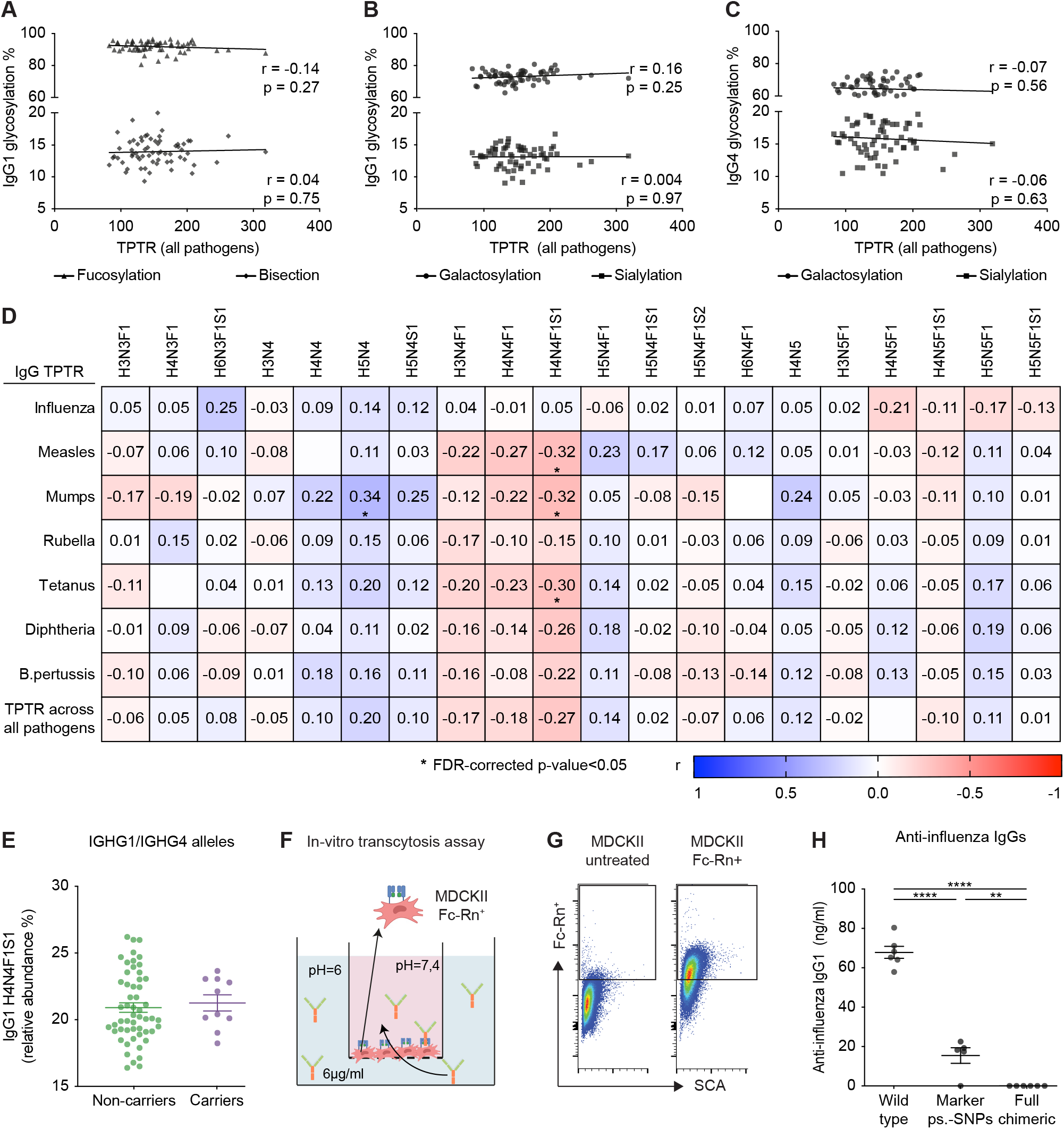
Functional analysis of chimeric IgG1/IgG4 antibodies on FcRn-mediated transcytosis and IgG Fc glycosylation. (**A-B**) Fucosylation, galactosylation, bisection, and sialylation of bulk IgG1 correlated with TPTR across all pathogens. (**C**) Galactosylation and sialylation of bulk IgG4 correlated with TPTR across all pathogens. (A-C) Pearson correlation was conducted. (**D**) Correlation matrix showing the relative abundance of glycans detected on IgG1, correlated with the TPTR of antibodies against specific pathogens and the overall TPTR across all pathogens. The Pearson correlation coefficient (r) is displayed in the boxes and color-coded according to the legend at the bottom graph. *;adjusted p value <0.05 (False Discovery Rate (FDR)-controlled by Benjamini-Hochberg procedure due to the high number of correlations in the dataset). Abbreviations used: H; hexose (mannose and galactose), N; N-acetylglucosamine, F; fucose, S; N-acetylneuraminic acid (sialic acid) (**E**). Relative abundance of the H4N4F1S1 glycan on bulk IgG1 among carriers and non-carriers of the IGHG1/IGHG4 alleles. IgG glycosylation analysis was performed using serum samples from the same mothers whose DNA was sequenced with Sanger sequencing. (**F**) Experimental design of the transcytosis assay using MDCKII FcRn^+^ cells, co-cultured with different biologically engineered IgG1 allotypes: a Wild-type IgG1, a construct harboring two duplication-marker pseudo-SNPs (Marker ps.-SNPs), and a construct carrying the full IgG1/IgG4 chimeric antibody sequence (Full chimeric) (table S8). (**G**) Representative flow cytometry plots confirming the successful transfection of MDCKII with the FcRn, using an APC-conjugated anti-alfa-tag nanobody. (**H**) Transcytosis of the respective biologically engineered anti-influenza IgG described in (F). Results are representative of two independent experiments, each with three replicates per IgG1 allotype. *;p < 0.05, **;p < 0.01, ***;p < 0.001, ****;p < 0.0001, determined by one-way ANOVA with multiple comparisons.

To further evaluate the relevance of the chimeric *IGHG1/IGHG4* duplications functionally, we conducted an *in vitro* transcytosis assay using polarized MDCKII cells stably expressing the FcRn receptor, to which we added three engineered IgG1/IgG4 chimera-like allotypes of a human anti-influenza IgG1 antibody (Fig. 4F,G, fig. S11) (*28*): 1) a wild-type IgG1; 2) a construct containing two duplication-marker pseudo-SNPs; and 3) a construct with all identified IgG1/IgG4 chimeric differences compared to canonical *IGHG1* (table S8). We observed significantly higher transcytosis in cultures with wild-type IgG1 compared to the full chimeric IgG1/IgG4 (Fig. 4H).

### Infants born to mothers with genetic duplications resulting in chimeric IGHG1/IGHG4 alleles have a greater risk of early-life respiratory infections

Next, we evaluated whether the different maternal allotypes affect the infants’ risk of infections during the first six and 12 months of life. (Fig. 5A). We identified that infants born to mothers carrying chimeric IGHG1/IGHG4 alleles exhibited a higher number of respiratory infections during the first six months of life compared to infants born to non-carrier mothers (Fig. 5B). Specifically, 80% of infants born to mothers carrying duplications leading to chimeric IGHG1/IGHG4 alleles experienced two respiratory infections in the first six months of life (Fig. 5C), with a relative risk of having 2 infections instead of 2.9 (Fig. 5C). By contrast, approximately 45% of infants born to mothers without this duplication experienced two infections within the same timeframe. The infection risk at 7-12 months, when passive immunity has waned, was unaffected by the presence or absence of maternal chimeric IGHG1 (Fig. 5D). These findings suggest a direct functional link between maternal IgG allotype architecture and early-life susceptibility to respiratory infections, highlighting how antibody levels and likely also the stability, and functionality of passively acquired maternal antibodies modulate infants’ health.

**Fig. 5.**
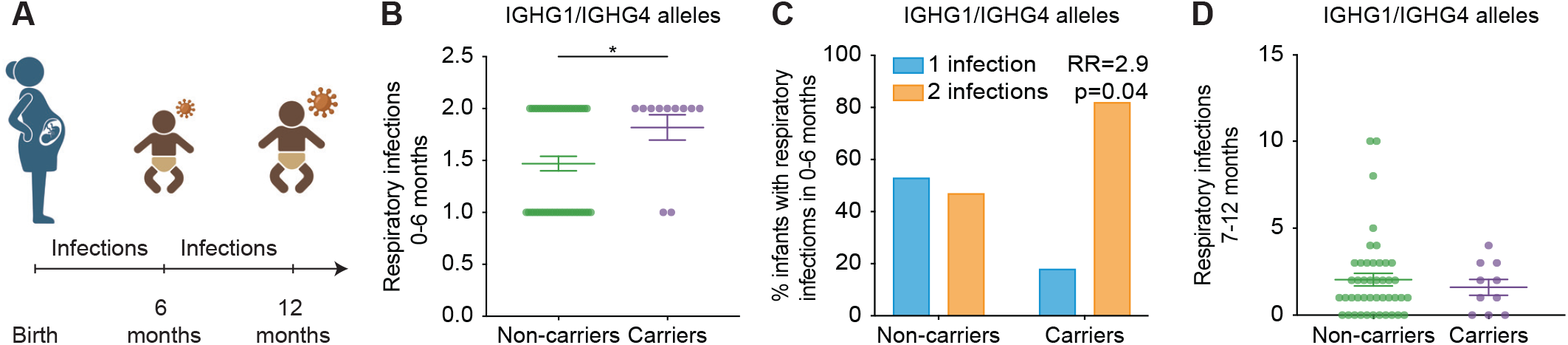
Infection in infants born to mothers carrying *IGHG1/IGHG4* duplications. (**A**) Schematic illustration of the study timeline, the number (N) of respiratory infections was documented at six months of life, followed by the assessment of respiratory infections between 7-12 months. (**B**) Respiratory infection rates during the first six months of life, stratified by maternal carriers and non-carriers of the chimeric *IGHG1/IGHG4* alleles. Data are shown as mean ± SEM. *;p < 0.05, as assessed by Welch’s t test. (**C**) Percentage of infants with one or two respiratory infections during the first six months of life, stratified by maternal carriers and non-carriers of the chimeric *IGHG1/IGHG4* alleles. Relative risk ratio (RR) of infants born to mothers carrying the *IGHG1/IGHG4* alleles to have two against one respiratory infection during the first 6 months of life, as assessed by Fisher’s exact test. (**D**) same as (C) in children during the 7-12 month postnatal period. Welch’s t-test was used to detect levels of significance.

## Discussion

Our research uncovers a previously unrecognized layer of a highly prevalent genetic variation in the maternal IGHG locus that fundamentally determines passive immunity in infants. We observed that over 20% of the pregnant study participants we analyzed carry a complex genetic IGHG rearrangement resulting in chimeric *IGHG1/IGHG4* genes. This variation leads to structural variants of the IgG1 antibody, which markedly reduce the efficiency of antibody transfer across the placenta, rendering infants more vulnerable to respiratory infections during the critical first six months of life.

This finding challenges the traditional view that passive immunity mainly depends on maternal antibody levels, glycosylation, or placental health. Instead, it emphasizes how the genetic structure of the antibody gene loci, especially when leading to structural variants, impacts infant immunity.

Notably, structural variants such as the IGHG1/IGHG4 duplication are often overlooked by standard genomic analysis because they are missing from current human reference genomes like GRCh38. This underscores a significant gap in both standard genomic research and routine diagnostic bioinformatics pipelines (*19*). Detecting such structurally complex yet functionally significant variants necessitates *de novo* assembly of long-read sequencing data (*29, 30*). Strikingly, despite their cryptic nature in typical next-generation sequencing analyses, we demonstrate that these variants can be reliably detected through Sanger sequencing by exploiting characteristic pseudo-SNP patterns arising from the underlying duplications. This offers a simple, cost-effective strategy to screen for clinically impactful IGHG structural variants.

Mechanistically, the decreased transfer of antibodies encoded by chimeric *IGHG1/IGHG4* genes—also observed in our in vitro transcytosis assay—is likely due to the IgG4-like properties, which are associated with reduced transplacental transport. (*11*).

Our findings not only impact maternal-fetal immunity but also have broader implications in immunogenetics, as we demonstrate that a functionally relevant structural variation in the IGHG gene occurs at an unexpectedly high frequency within the European population (*19, 20*). Standard methods that mainly catalogue SNPs might overlook such essential insights of antibody diversity. Since immunoglobulin structure is essential for responses to infections, vaccines, autoimmunity, and therapeutic antibodies, recognizing these hidden variations is important for gaining a deeper understanding of individual immune differences.

The unexpectedly high frequency of these chimeric *IGHG1/IGHG4* genes and their negative impact on infants’ protection from infections pose significant evolutionary questions. (*31, 32*). One possible explanation is that their IgG4-like characteristics could enhance maternal-fetal immune tolerance by diminishing Fc-mediated inflammatory responses at the placental interface, which may support implantation or reduce the risk of pregnancy complications (*33*–*35*). Variants that enhance reproductive success whilst rendering offspring more susceptible to diseases have been proposed as a classic example of an evolutionary trade-off (*36*).

Our insights hold considerable clinical importance. Infections in early infancy continue to be a major health concern, and in Western countries, the expenses linked to hospitalization due to infant infections are considerable (*2, 37*). While vaccination during pregnancy can mitigate the risk of early-life infections, compliance with the current guidelines remains low (*38, 39*). Our study’s findings strongly underpin that passive immunity can vary even among healthy pregnant individuals due to previously unknown but prevalent maternal genetic factors. This further highlights the need for individualized vaccination strategies during pregnancy, which might enhance protection for newborns.

In summary, we identify previously unrecognized genetic variants in mothers that impair passive immunity of infants. Additionally, we introduce a straightforward surrogate test to detect these variants. Applying this method in clinical settings will enable personalized vaccine strategies and support the creation of customized vaccines, ultimately enhancing protection for both mothers and their babies.

## Data Availability

All data produced in the present study are available upon reasonable request to the authors

## Acknowledgments

The authors thank all PRINCE participants for permitting their study during pregnancy and for their ongoing cooperation. They also appreciate Gudula Hansen’s support in managing subject recruitment and Lina Otto’s assistance with database queries

## Funding

German Center for Child and Adolescent Health, Hamburg site (PCA, AD)

Federal Ministry of Research and Education: Junior Research Center on Reproduction (PCA, AD)

German Research Foundation: CRC 1713, 91232/1-1713 (PCA, AD), KFO296: DI2103/2-2 (AD), AR232/24-2 (PCA), RU5068: AR232/29-2 (PCA), SCHL2276/2-1 (CS), TRR333/1, 450149205 (CS)

Next Generation Partnership, Excellence Initiative University Hamburg (PCA, AD)

State Ministry of Research and Education and Equality (LFF-FV73) (PCA, AD)

Werner Otto Foundation (DEZ)

Leibniz Science Campus InterACt (supported by the BWFGB Hamburg and the Leibniz Association) (KMC, MT)

## Author contributions

Conceptualization: PCA, CS, AD.

Methodology: IB, IE, JH, TS, KR, MA, CM, MS, AW, KR, KT, AG, IG, CU, SGZ, WW, SG, JN, KMC, MT, FS, FKN, ML, MW, MA, MB, MMA, DEZ, AD, CS

Investigation: IB, IE, JH, KR, MA, AW, KR, WW, SG, JN

Software: TS, CM, MS, MB, MA, IB

Validation: IB, IE, JH, KR, TS, CM, MS

Visualization: IB, CS, IE, TS, CM, MS, MB, MA, KMC, MT

Formal analysis: IB, CS, IE, TS, CM, MS, MB, MA, WW, SG, JN, KMC, MT

Resources: IB, IE, JH, TS, KR, MA, CM, MS, AW, KR, KT, AG, SGZ, IG, CU, WW, SG, JN, KMC, MT, FS, FKN, ML, MW, MA, MB, MMA, DEZ, AD, CS, PCA

Funding acquisition: PCA, CS, AD, DZ

Project administration: PCA, IB, CS, AD

Supervision: PCA, CS

Writing – original draft: IB, CS, PCA

Writing – review & editing: IB, IE, JH, TS, KR, MA, CM, MS, KT, AG, SGZ, IG, CU, WW, SG, JN, KMC, MT, FS, FKN, ML, MW, MA, MB, MMA, DEZ, AD, CS, PCA

## Competing interests

The Authors declare that they have no competing interests.

## Data and materials availability

All data, code, and materials used in the analysis are available upon request to any researcher for purposes of reproducing or extending the analysis.

## Supplementary Materials

Materials and Methods

Figs. S1 to S11

Tables S1 to S8

## Notes

### Competing Interest Statement

The authors have declared no competing interest.

### Author Declarations

All study participants provided signed informed consent forms. The study protocol was approved by the ethics committee of the Hamburg Chamber of Physicians (license number PV 3694). It was conducted in compliance with the Declaration of Helsinki for Medical Research involving Human Subjects.

