## Supplement for "Maternal *IGHG* locus duplications impair infants’ passive immunity"

**Affiliations:**

**The PDF file includes:**

Materials and Methods

Figs. S1 to S11

Tables S1 to S8

**Materials and Methods**

**Study subjects**

Our study is embedded in the PRINCE (Prenatal Identification of Children’s Health) cohort, a longitudinal, prospectively-designed pregnancy cohort study taking place at the University Medical Center Hamburg-Eppendorf since 2011 (n=780). Inclusion criteria included age ≥ 18 years and a viable pregnancy of 12–14 weeks of gestation. Exclusion criteria were chronic infections (HIV, hepatitis B or C), drug or alcohol abuse, multiple gestations or pregnancies deriving from assisted reproductive technology. Pregnancy progression, maternal health status, medication, stress perception, and anthropometric data were documented at three time points between 12-14, 22-24, and 34-36 gestational weeks. During these study visits, transabdominal ultrasound and maternal blood sample collection were also performed. At birth, neonatal anthropometric parameters were documented, and cord blood was collected. After birth, documentation of the children’s health status, including the number and type of infections, continued annually. Among all PRINCE study participants, n=139 women participated with at least two subsequent pregnancies . We here focused on PRINCE participants with two consecutive pregnancies, for which complete clinical information as well as maternal and cord serum samples were available (n=38 mothers with a total of 76 children born). Relevant demographic characteristics are shown in Table S1. With regard to the exome sequencing of the *IGHG1* gene, we worked with a validation subset of the PRINCE study, for which complete clinical information, maternal and cord blood serum, as well as maternal whole blood samples were available (n=66 pregnant women).

**Study approval**

All study participants provided signed informed consent forms. The study protocol was approved by the ethics committee of the Hamburg Chamber of Physicians (license number PV 3694). It was conducted in compliance with the Declaration of Helsinki for Medical Research involving Human Subjects.

**Blood samples and infection information**

During each gestational study visit, a venous blood sample was obtained from the mother by peripheral venipuncture. At birth, cord blood was taken from the umbilical cord after cord clamping. Serum samples were stored at -80°C until use and were kept at 4°C after thawing. We exclusively used maternal samples taken between gestational week 22 and 24. The documentation of the infection incidence during the first six and twelve months of the infant’s life was based on standardized parental questionnaires and complemented by documentation of the child’s pediatrician. Reported respiratory infections included common cold, tonsillitis, croup, bronchitis, and pneumonia.

**Enzyme-linked immunosorbent assays (ELISAs)**

Using commercially available EUROIMMUN ELISA kits, we measured IgG antibody titers against eight vaccine-preventable pathogens in maternal serum collected between 22 and 24 weeks of gestation, as well as in corresponding cord blood serum. The targeted IgG antibodies included anti-Tetanus Toxoid, anti-Diphtheria Toxoid, anti-Influenza A virus, anti-Bordetella pertussis toxin, anti-RSV, anti-Rubella, anti-Measles, and anti-Mumps. Assays followed the manufacturer’s protocols. Influenza, RSV, and Mumps titers were expressed in Relative Units per milliliter (RU/ml), while Tetanus, B. pertussis, Diphtheria, and Rubella titers were in International Units per milliliter (IU/ml). Measles titers were measured in IU/l. Samples with antibody levels above the detection threshold were deemed seropositive. Due to limited serum volume and handling procedures, not all titers could be assessed in every sample. All tests were performed in duplicates. Maternal and cord blood samples from the same pregnancy were processed on the same microtiter plate to minimize inter-assay variation. For measuring the biologically engineered IgG1 allotypes used in the transcytosis assay, a commercially available human IgG1 ELISA Kit (Invitrogen) was utilized.

**Assessment of the transplacental antibody transfer rate**

The TPTR percentage was calculated using the formula: (cord blood IgG concentration / maternal IgG concentration) * 100. For each pathogen-specific antibody, the mean TPTR across all individuals in both first and second pregnancies was computed, and then the deviation from this mean was determined for each pregnant woman, resulting in TPTR z-scores. To identify women with consistently higher TPTR across all pathogens and both pregnancies, we averaged the TPTR z-scores for all antibodies per individual and divided these into quartiles. The lowest quartile (bottom 25%) was defined as low TPTR individuals (n=9), while the highest quartile (top 25%) was classified as high TPTR individuals (n=8). The middle two quartiles were considered moderate TPTR individuals (n=21). The average TPTR for both pregnancies was then calculated for each individual within these quartiles. For the validation subset of the study, the average of all TPTR values for each individual was calculated, covering antibodies against Tetanus, Diphtheria, Influenza A virus, Bordetella pertussis, Rubella, Measles, and Mumps (TPTR across all pathogens).

**Sanger sequencing**

DNA was extracted from whole blood samples of 66 pregnant women participating in the PRINCE study. IGHG1 screening was conducted using standard PCR amplification and Sanger sequencing techniques. For *IGHG1* sequencing, established primer sequences were employed (*1*). All exons and flanking intronic regions were compared to reference sequences (*IGHG1*: ENST00000631539.1, *IGHG2*: ENST00000641095.1, *IGHG3*: ENST00000641136.1, *IGHG4*: ENST00000641978.1). For Sanger haplotyping, PCR amplicons were cloned into vectors with the TOPO TA Cloning Kit pCR 2.1-TOPO (Invitrogen), transformed into TOP10 competent cells (ThermoFisher), and DNA from individual clones was sequenced following standard protocols.

**Long-read sequencing and analysis**

HiFi long-read sequencing was performed in five individuals with Sanger-confirmed complex haplotypes. Genomic DNA was extracted from blood samples, and DNA quality control was carried out by standard fluorescence-based quantification methods. One sample failed quality control due to insufficient coverage of the *IGHG* locus (coverage <5x). For the remaining four, Library preparation was performed using the SMRTbell prep kit 3.0 (PacBio), and sequencing was carried out using a PacBio Revio system. Demultiplexing of the sequencing reads was executed with PacBio lima (2.12.0). HiFi reads (CCS reads with a predicted accuracy ≥ Q20) were extracted using SAMtools (1.21) (*2*) and converted to FASTQ format using PacBio bam2fastq (1.4.0). Sequence reads were assembled into contigs using Flye (*3*) (v2.9.6-b1802) with the additional parameter '--pacbio-hifi'. To align primer sequences to the contigs, GLSEARCH (*4*) (v36.3.8i) was employed with the parameters '-n -m 8C'. The results were saved in a commented BLAST tabular format and subsequently parsed to identify primer pairs that aligned within a maximum distance of 5 kb from each other. For each putative amplicon identified, SAMtools (*5*) faidx (v1.21) was used to extract the corresponding sequence from the respective contig.

**Structural analysis of IgG1 SNPs**

We analyzed the structure of the chimeric IgG1/IgG4 and its FcRn binding site by mapping the SNPs onto the IgG1 structure. For this, we used three X-ray structures from the Protein Data Bank (PDB) (*6*): one of IgG1 bound to FcRn (PDB ID 4N0U) (*7*), another of IgG1 bound to influenza hemagglutinin (PDB ID 5UGY) (*8*), and a third showing the interaction between the two heavy constant chains in an IgG1 molecule (PDB ID: 5JII) (*9*). All visualizations were carried out using UCSF ChimeraX, version 1.9 (*10*). Interface analyses were supported by the PICKLUSTER Chimera plug-in (*11*).

**IgG Fc glycosylation analysis – “GLYcoLISA”**

Bulk IgG was extracted from serum samples of pregnant participants in the PRINCE study through protein G affinity purification, tryptic digestion, and LC–MS analysis of tryptic IgG Fc glycopeptides. This process included calculating galactosylation, sialylation, bisection, and fucosylation, as previously described (*12*).

**Production of biologically engineered anti-influenza IgG1 allotypes**

Three human monoclonal IgG1 allotypes of the same anti-influenza antibody were produced by GenScript (Nanjing, China) using the sequence from a previously characterized human anti-influenza IgG1 antibody (*13*). The low-mutated IgG1 allotype was created based on the wild-type (wt) anti-influenza IgG1 sequence, representing individuals in the cluster with low SNP frequency. The moderate-mutated IgG1 allotype was generated by introducing changes caused by rs1063407 and rs587637831 (the missense TPTR_low_ SNPs), reflecting individuals with moderate SNP variation. The highly mutated IgG1 allotype incorporated all the missense SNPs found in participants with the highest SNP frequency. All SNPs used in engineering the antibodies were assumed to be homozygous, ensuring the mutations were expressed. Specific amino acid sequences for each allotype are detailed in table S8.

**Transcytosis assay**

Lentiviral vectors, one encoding the N-terminally ALFA-tagged full-length human Fc-Rn and another encoding N-terminally FLAG-tagged human β2-microglobulin, were produced in 293T cells, as previously described (*14*, *15*). MDCKII cells were sequentially transduced with both vectors to express human Fc-Rn and β2-microglobulin. MDCKII Fc-Rn+ cells were isolated via puromycin selection and expanded. These cells were then compared to untreated MDCKII cells by flow cytometry using an APC-conjugated anti-ALFA-tag nanobody (*16*) to assess Fc-Rn expression. Cells were seeded at a density of 50000 cells per Transwell filter (optimized as in figure S11E). After 72 hours, the medium was refreshed, and a transcytosis assay was conducted as previously described (*17*). TEER measurements of each Transwell were taken daily to confirm the formation of a fully confluent and polarized monolayer. A concentration of 6 µg/mL of each IgG1 allotype was added to the basal compartment of the Transwells. After 2 hours, IgG1 allotype levels in the apical medium were quantified by ELISA. The medium in the basal compartment was adjusted to pH 6 with Minimal Essential Medium (MEM), while the apical medium remained at pH 7.4. Following incubation, the upper compartment medium was collected for IgG1 allotype measurement using the specified human IgG1 ELISA kit.

**Statistical/bioinformatical analysis**

Maternal and neonatal demographic and anthropometric parameters, along with all antibody titer values, are shown in the tables as mean ± SD. Continuous data comparisons between two groups were conducted using paired t-test, Mann-Whitney, or Welch’s test, based on the data distribution and variance. For comparisons across more than two groups, one-way ANOVA with multiple comparisons was used. Categorical data were analyzed with Chi-square or Fisher’s Exact test. Antibody titer comparisons employed paired t-test and Wilcoxon test, depending on the data distribution, with a significance threshold of p ≤ 0.05. Dot plot graphs display means ± SEM. Analyses and plots were generated using GraphPad Prism version 9.5.1 (GraphPad Software, Inc., La Jolla, CA), R version 4.1.2, and Python 3.8. Icon-based figures were created with BioRender.com.

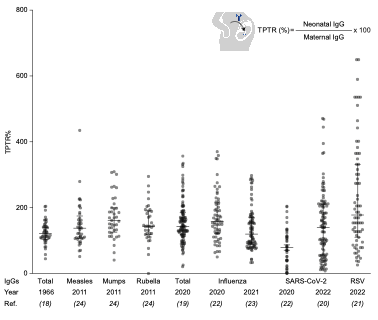

Fig. S1: Variation in transplacental antibody transfer rate (TPTR) across different studies published in the past. The dot plot shows the range of TPTR in mother/infant (cord blood) pairs, based on data from studies (*18*–*24*). The top right corner features the equation used to calculate TPTR.

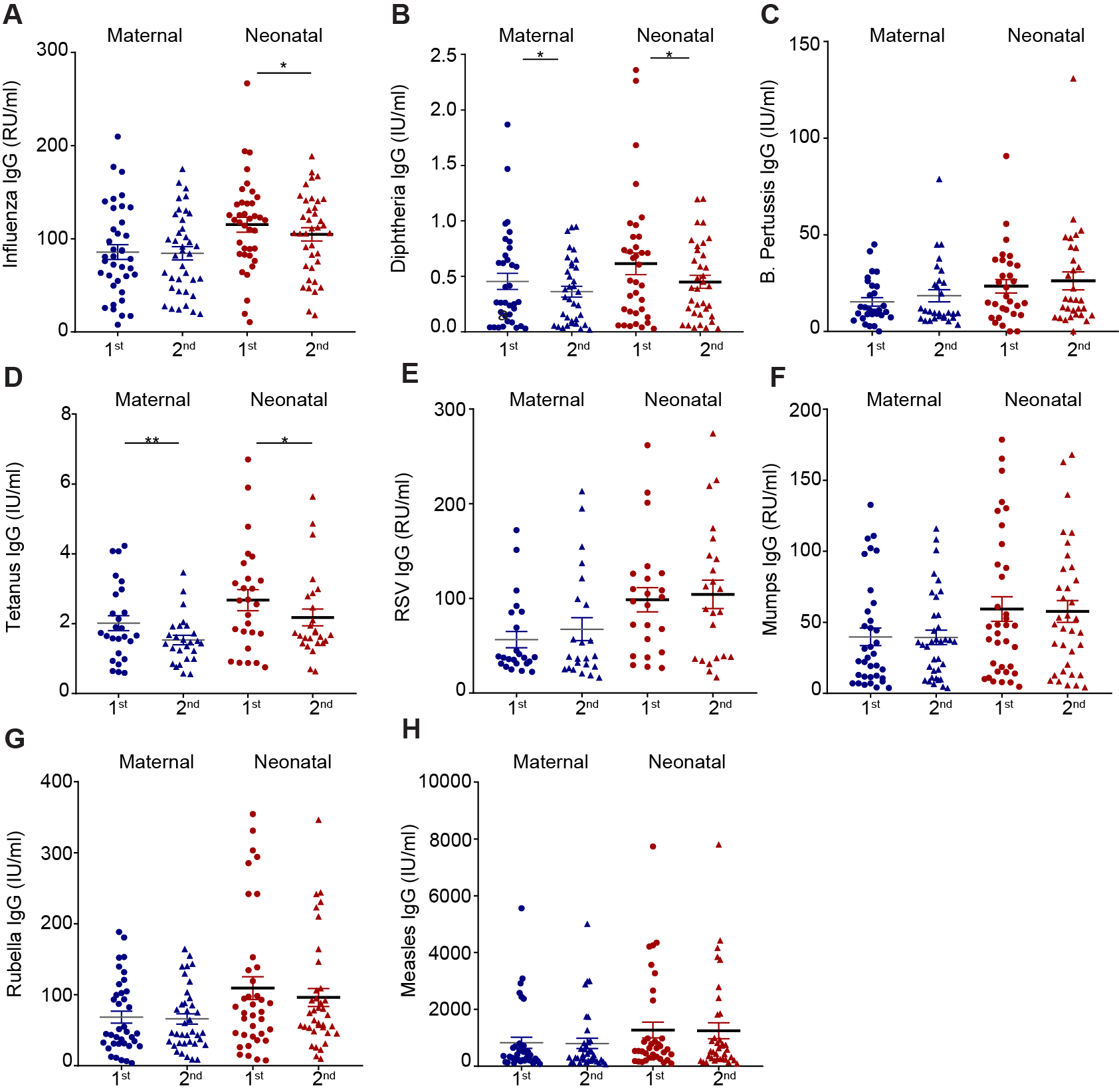

Fig. S2: Maternal and neonatal (cord blood) IgG antibody levels during first and second pregnancies. Maternal and neonatal IgG titers are depicted for (A) Influenza, (B) Diphtheria, (C) B. pertussis, (D) Tetanus, (E) RSV, (F) Mumps, (G) Rubella, and (H) Measles, comparing the two pregnancies. Data are presented as mean ± SEM. Significant differences are indicated by * (p < 0.05) and ** (p < 0.01), based on paired t-test or Wilcoxon test, depending on data distribution. Differences with p > 0.05 are considered non-significant and are not explicitly marked.

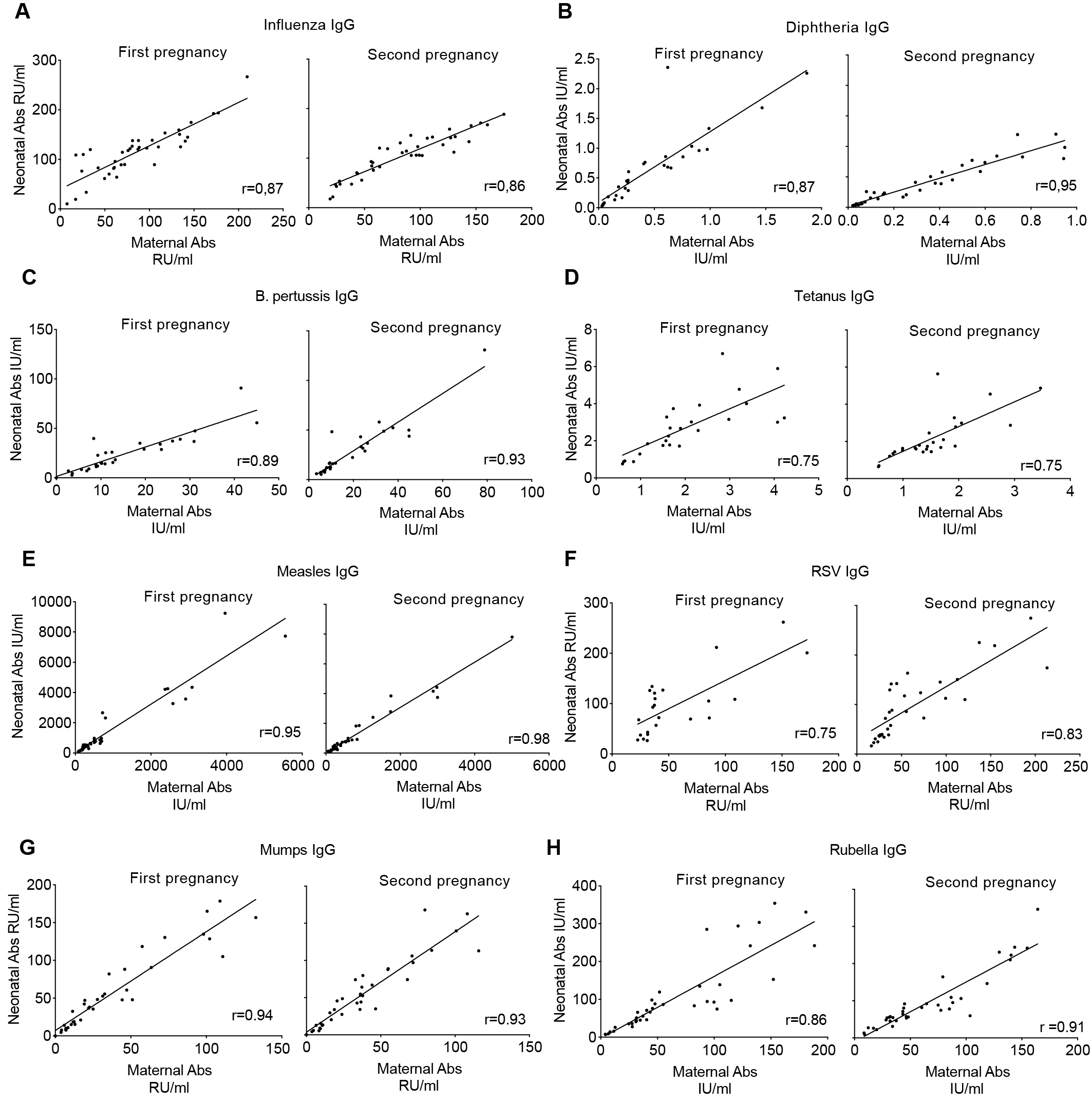

Fig. S3: Correlation between maternal and cord blood IgG serum levels in first and second pregnancies. Maternal-neonatal (cord blood) correlation of (A) Influenza, (B) Diphtheria, (C) B. pertussis, (D) Tetanus, (E) Measles, (F) RSV, (G) Mumps, and (H) Rubella IgG antibodies in the first and second pregnancy. Each dot represents a mother/neonate pair. All correlations are significant with p < 0.001, as calculated using the Pearson Correlation test. Pearson’s correlation coefficient (r) is shown on each graph.

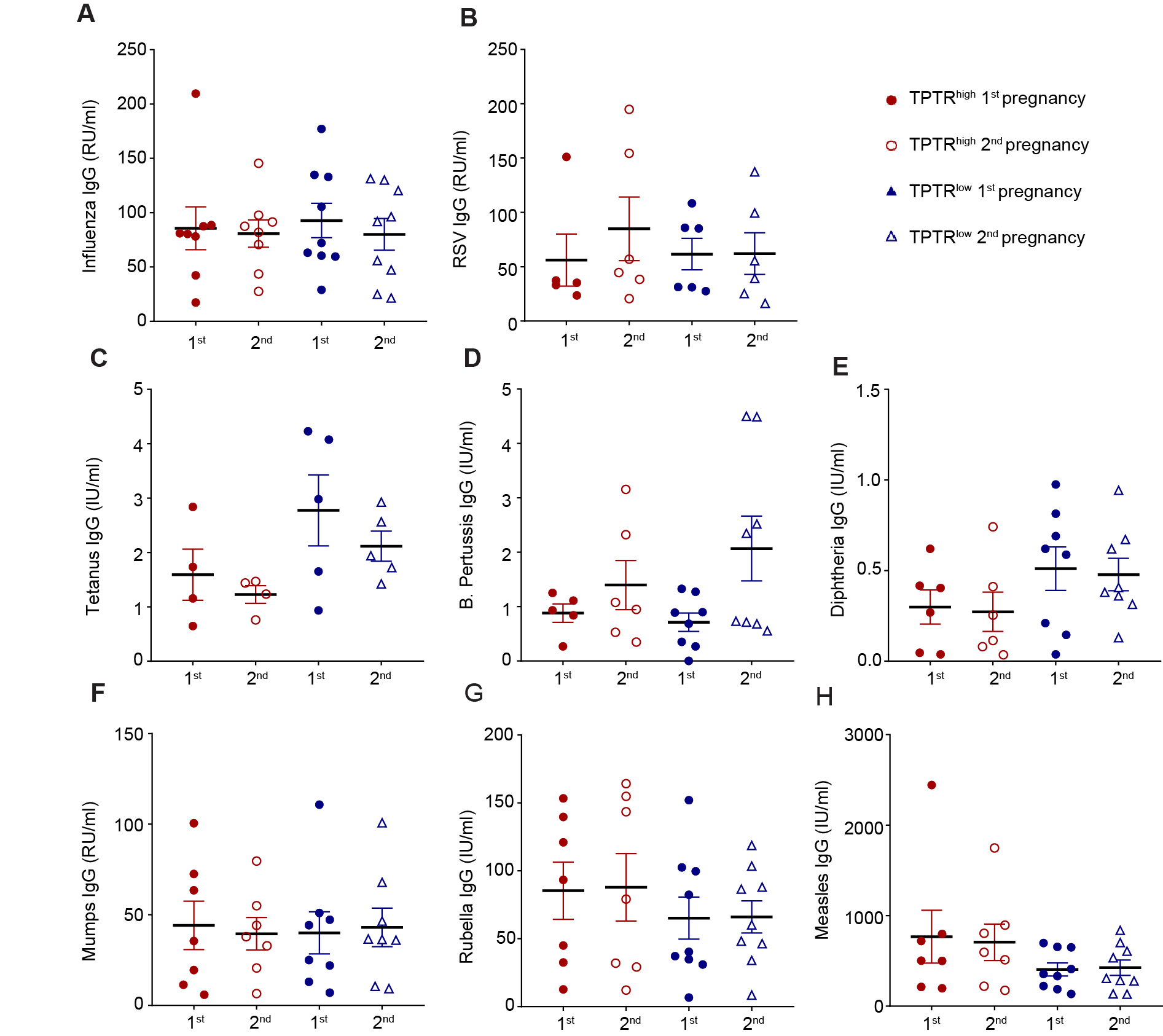

**Fig. S4: Comparison of serum IgG antibody titers during the first and second pregnancies in TPTR^high^ and TPTR^low^ mothers.** Pathogen-specific IgG against **(**A) Influenza, (B) RSV, (C) Tetanus, (D) B. Pertussis, (E) Diphtheria, (F) Mumps, (G) Rubella, and (H) Measles in first and second pregnancies. Data are shown as mean ± SEM. No statistically significant differences were found, as determined by paired t-test or Wilcoxon test, depending on data distribution.

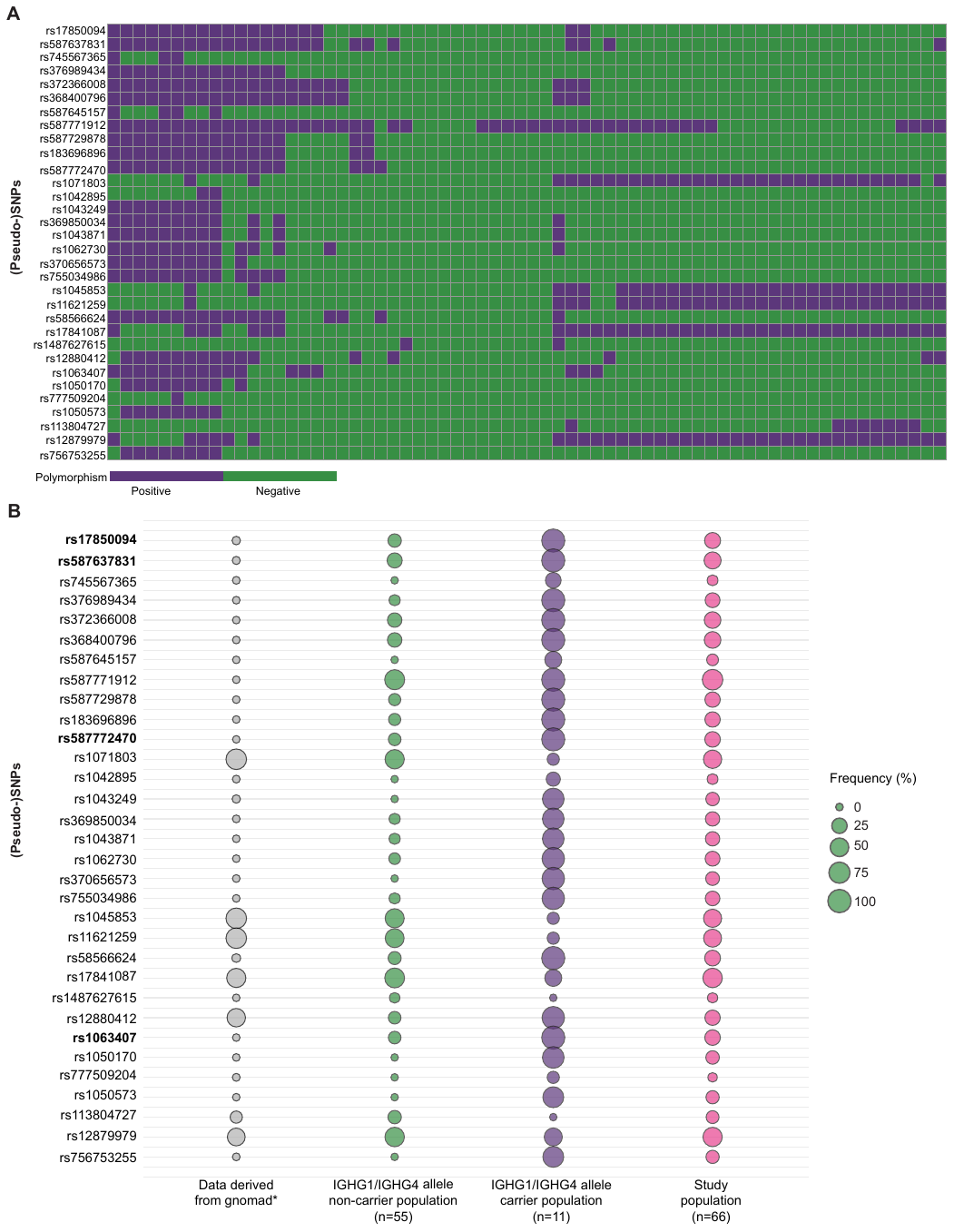

Fig. S5: Sanger-based detection of (pseudo-)SNPs across study individuals. (A) Heat map showing the main detectability of each (pseudo-)SNP at the genomic level in all individuals. (B) Bubble plot displaying the frequency of these pseudo-SNPs in different populations. The Gnomad database was used to compare our findings with publicly available data. *Note that for all SNPs identified here, the Gnomad database issues this warning: “This variant is covered in fewer than 50% of individuals in gnomAD v4.1.0 exomes. Allele frequency estimates may not be reliable.”

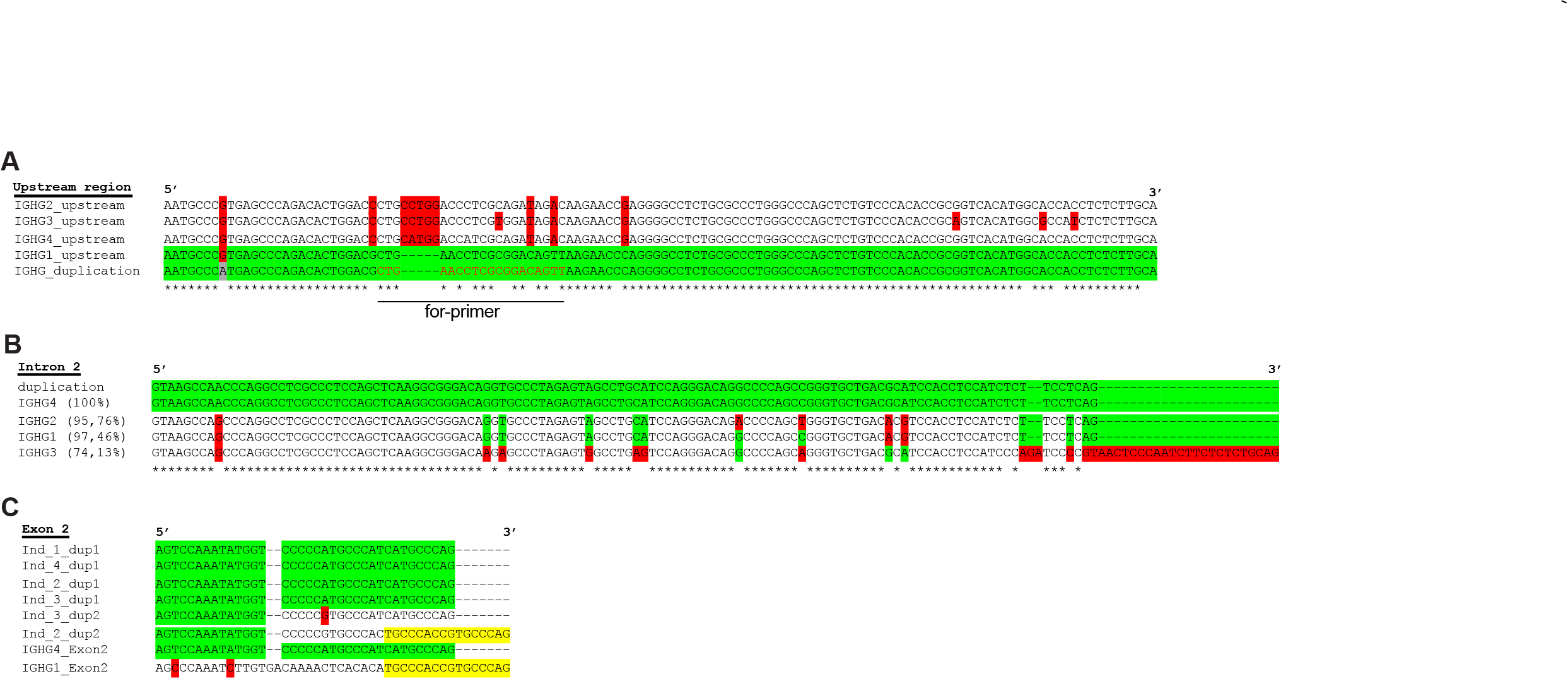

Fig. S6: Sequence alignments comparing duplicated alleles reveal chimeric *IGHG1/IGHG4* structures. (A) Multiple sequence alignment of upstream regions from canonical *IGHG* genes (*IGHG1-4*) and the duplication sequence shows that the 5’ upstream region of the duplication closely resembles *IGHG1* (green), suggesting that *IGHG1* regulatory elements influence the duplicated gene’s expression. Divergent nucleotides are marked in red. The primer binding site used in diagnostic Sanger sequencing is indicated below the alignment. (B) An example alignment of intron 2 sequences from the duplication and canonical *IGHG* genes shows that the duplicated allele most closely matches IGHG4, as seen in this intron 2 sample. Red marks indicate single nucleotide mismatches that differentiate it from *IGHG1* and other *IGHG* genes. (C) Alignment of exon 2 sequences from individuals with duplications (labeled Ind[.]) compared to reference *IGHG1* and *IGHG4* exon 2 sequences reveals a chimeric structure in one duplicated allele—comprising an *IGHG4*-like segment (green) at the 5’ end and an *IGHG1*-like segment (yellow) at the 3’ end, separated by a transitional region. Red highlights mismatch positions, confirming intra-exonic hybridization between *IGHG1* and *IGHG4* within the duplicated gene.

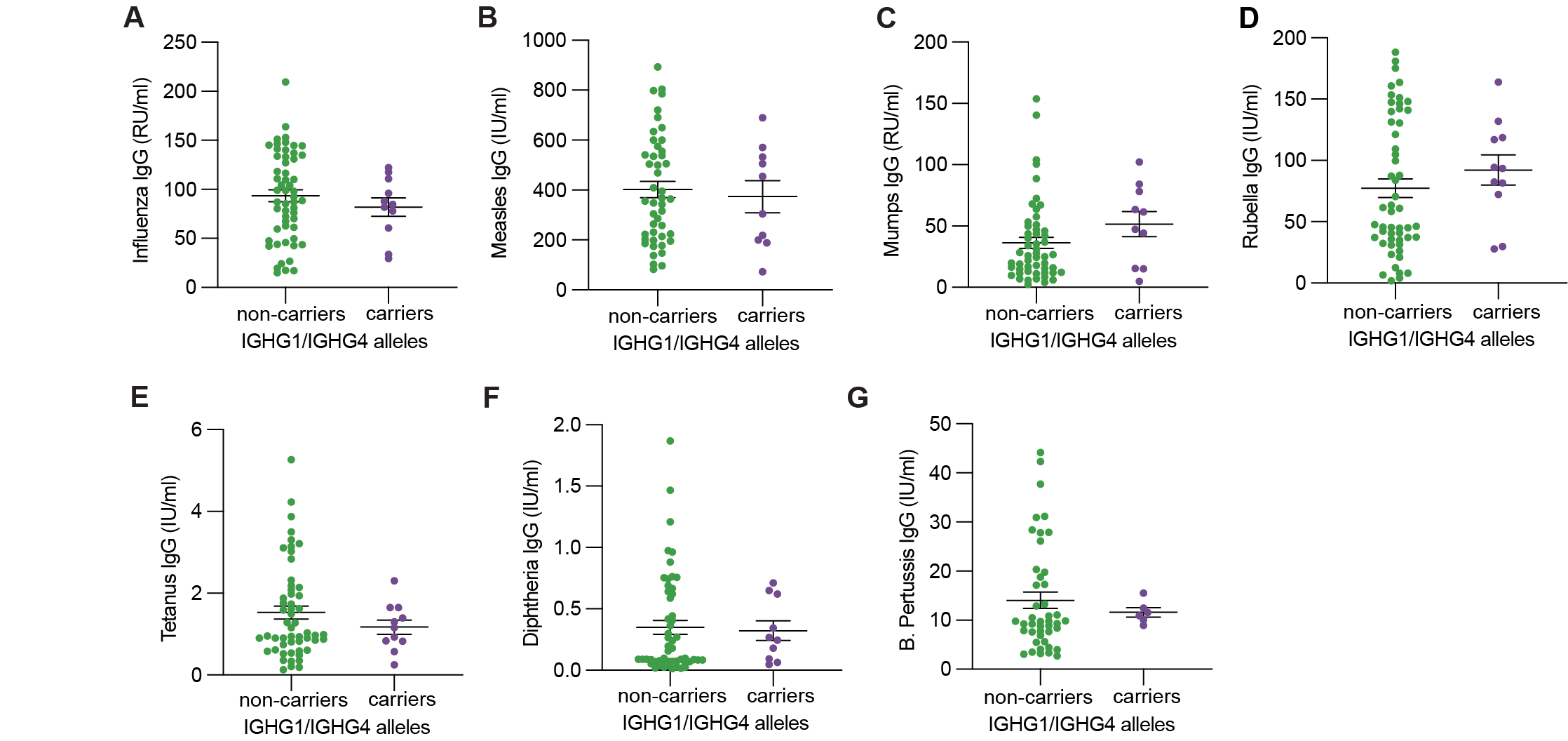

**Fig. S7: Serum antibody titers in pregnant mothers with or without chimeric IGHG1/IGHG4 alleles.** (A) Influenza, (B) Measles, (C) Mumps, (D) Rubella, (E) Tetanus, (F) Diphtheria, (G) B. Pertussis IgG antibodies are compared between those carrying the chimeric alleles and control mothers. Data are presented as mean ± SEM. No statistically significant differences were found, as determined by an unpaired t test.

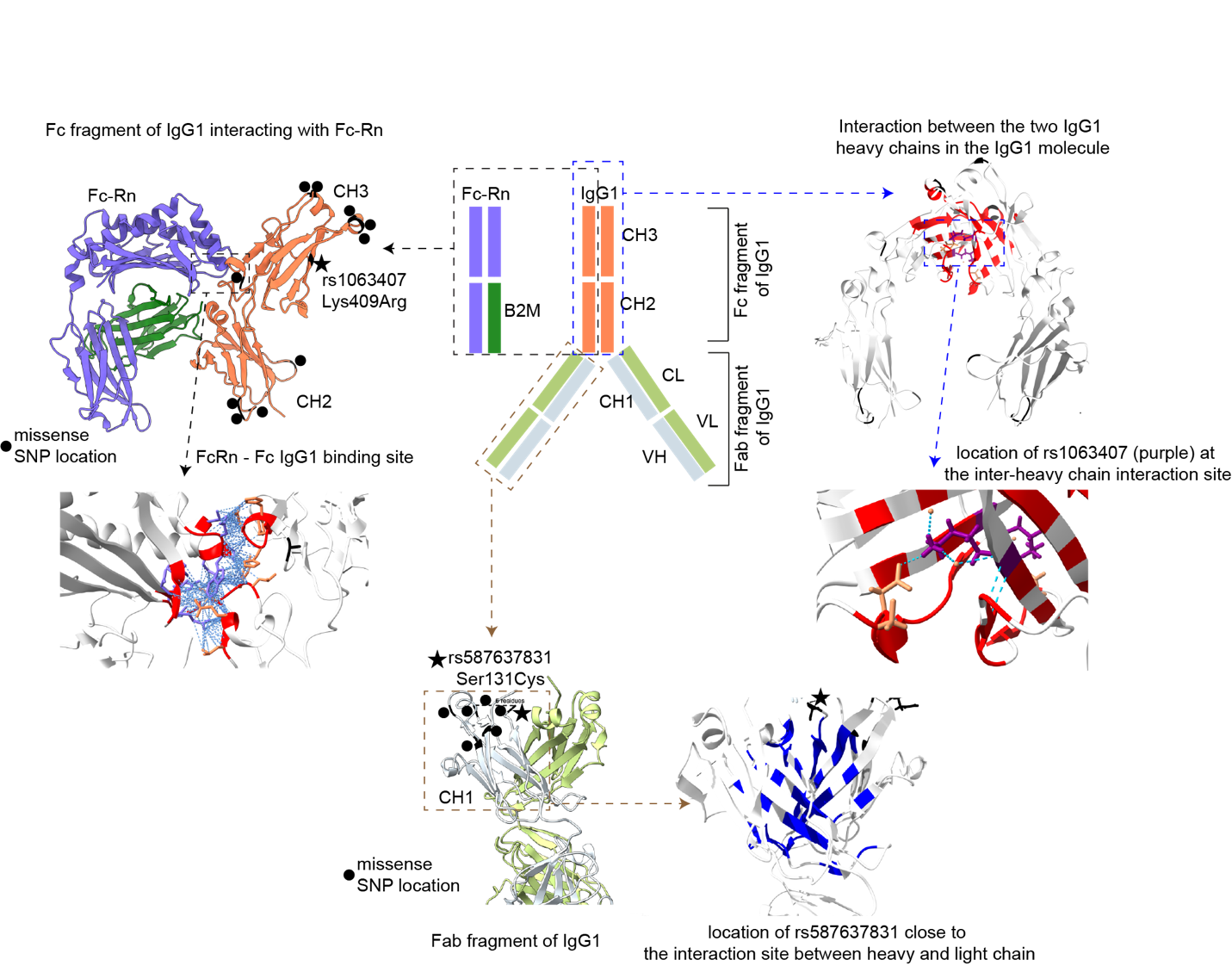
**Fig. S8: Structural modification from IgG1 to chimeric IgG1/IgG4.** Structural models of IgG1 and Fc-Rn are displayed, highlighting SNP locations. At the top center, the coloring scheme is shown. On the top left, the interaction between the IgG1 heavy chain and Fc-Rn (PDB ID: 4N0U) is depicted, with SNPs marked in black. The interaction surface is detailed below, with residues involved in contact highlighted in red. None of the SNPs are located at the IgG1-Fc-Rn interface. On the top right, the IgG1 heavy chain dimer (PDB ID: 5JII) is shown, along with a close-up of the interaction between heavy chains, with the contact area highlighted in red. The amino acid affected by rs1063407 appears in purple within the interaction surface and may influence the stability of the heavy chain dimer. Other SNPs are shown in black and are not within the interaction surface. At the bottom center, the structure of the anti-influenza IgG1 heavy and light chains is presented, with SNP locations marked by black dots. The black stars indicate the domains of two missense marker SNPs. In the zoomed-in view of the interaction surface (blue), one missense marker SNP is visible at the edge of the interface between IgG1 heavy and light chains, potentially affecting IgG1 stability, but likely to a lesser extent than rs1063407. Interface detection was performed using the tool PICKLUSTER(11) in all cases. Abbreviations include: Constant Heavy (CH) 1; the first constant domain of the IgG1 heavy chain, CH2; the second constant domain of the IgG1 heavy chain, CH3; the third constant domain of the IgG1 heavy chain, Variable Heavy (VH); the variable domain of the IgG1 heavy chain, Constant Light (CL); the constant domain of the IgG1 light chain, Variable Light (VL); the variable domain of the IgG1 light chain, Fragment antigen-binding (Fab), Fragment crystallizable (Fc region), B2M; b2-microglobulin. The lower panel illustrates the structure of anti-influenza IgG1 heavy and light chains, with black dots denoting pseudo-SNP locations. Black stars highlight the domains of the two missense marker pseudo-SNPs. In the detailed interaction surface view (blue), one missense marker pseudo-SNP appears at the edge of the interface between the IgG1 heavy and light chains, which might influence IgG1 stability, though likely to a much lesser degree than rs1063407. Interface detection was performed using the tool PICKLUSTER.

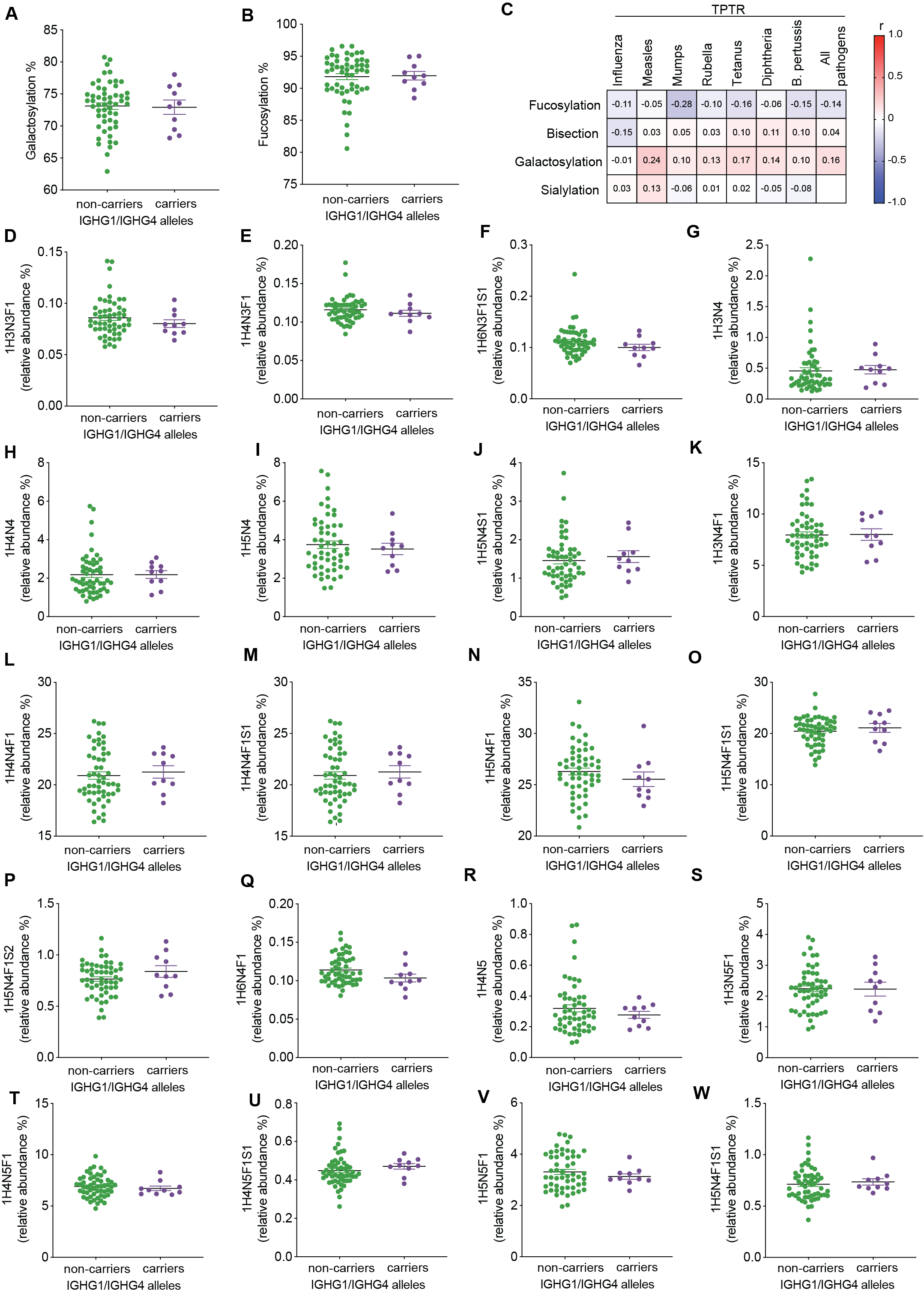

**Fig. S9: Bulk IgG1 Fc glycosylation shows no difference between pregnant women with or without chimeric IgG1/IgG4 antibodies.** (A-B) Galactosylation and fucosylation levels of bulk IgG1 in mothers with chimeric IgG1/IgG4 alleles compared to those without. Non-significant results are not highlighted, as determined by unpaired t-test. (C) Correlation matrix displaying the relative abundance of glycosylation features of IgG1, correlated with the TPTR of antibodies against specific pathogens and across all pathogens. The Pearson correlation coefficient (r) is shown in the boxes, with color coding to the right. Non-significant correlations (adjusted p-value > 0.05) are not highlighted, with FDR control applied via the Benjamini-Hochberg procedure due to multiple comparisons. (D-W) Relative abundance of specific glycans on bulk IgG1 among mothers with and without chimeric IgG1/IgG4. *; p-value < 0.05, as determined by unpaired t-test. Non-significant results are not highlighted.

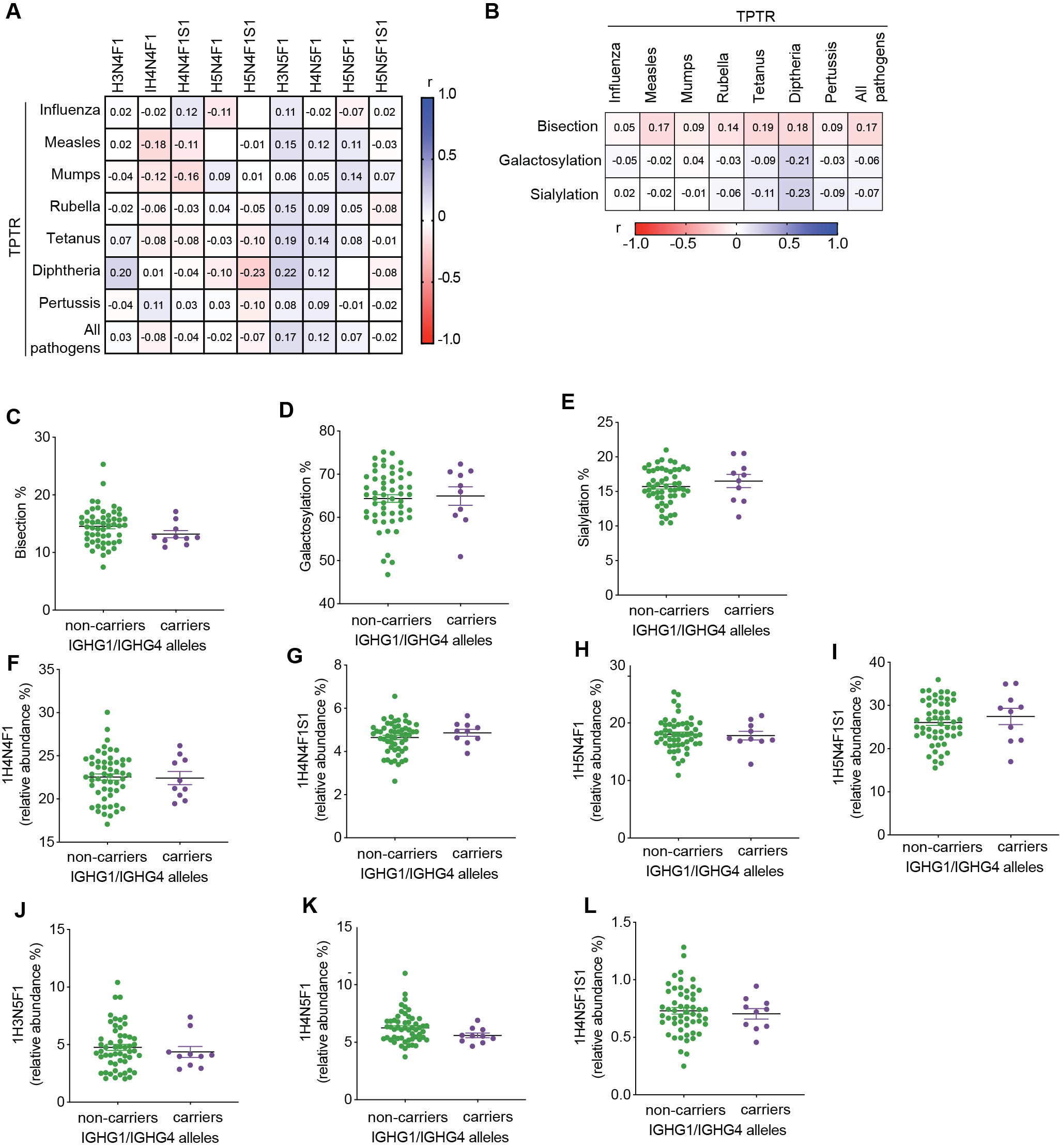

**Fig. S10: Bulk IgG4 Fc glycosylation shows no difference between pregnant women with or without chimeric IgG1/IgG4 antibodies.** (A) A correlation matrix displaying the relative abundance of glycans on IgG4 linked to the TPTR of antibodies against specific pathogens and the overall TPTR across all pathogens. Pearson correlation coefficients (r) are provided in the boxes and color-coded on the right. Non-significant differences (adjusted p-value > 0.05) are not highlighted, with FDR controlled by the Benjamini-Hochberg method due to numerous correlations. (B) A correlation matrix of IgG4 glycosylation features correlated with the TPTR of antibodies against specific pathogens and overall. Pearson r values are shown in the boxes with right-side color coding. *; indicates adjusted p-value < 0.05, FDR-controlled as above. (C) Bisection, (D) Galactosylation, and (E) Sialylation levels of bulk IgG4 among mothers with or without chimeric IgG1/IgG4 alleles. Non-significant results are not highlighted, as determined by unpaired t-test. (F-L) Relative abundance of specific glycans on bulk IgG4 in mothers with or without chimeric IgG1/IgG4 alleles. Non-significant findings are not highlighted, based on unpaired t-test.

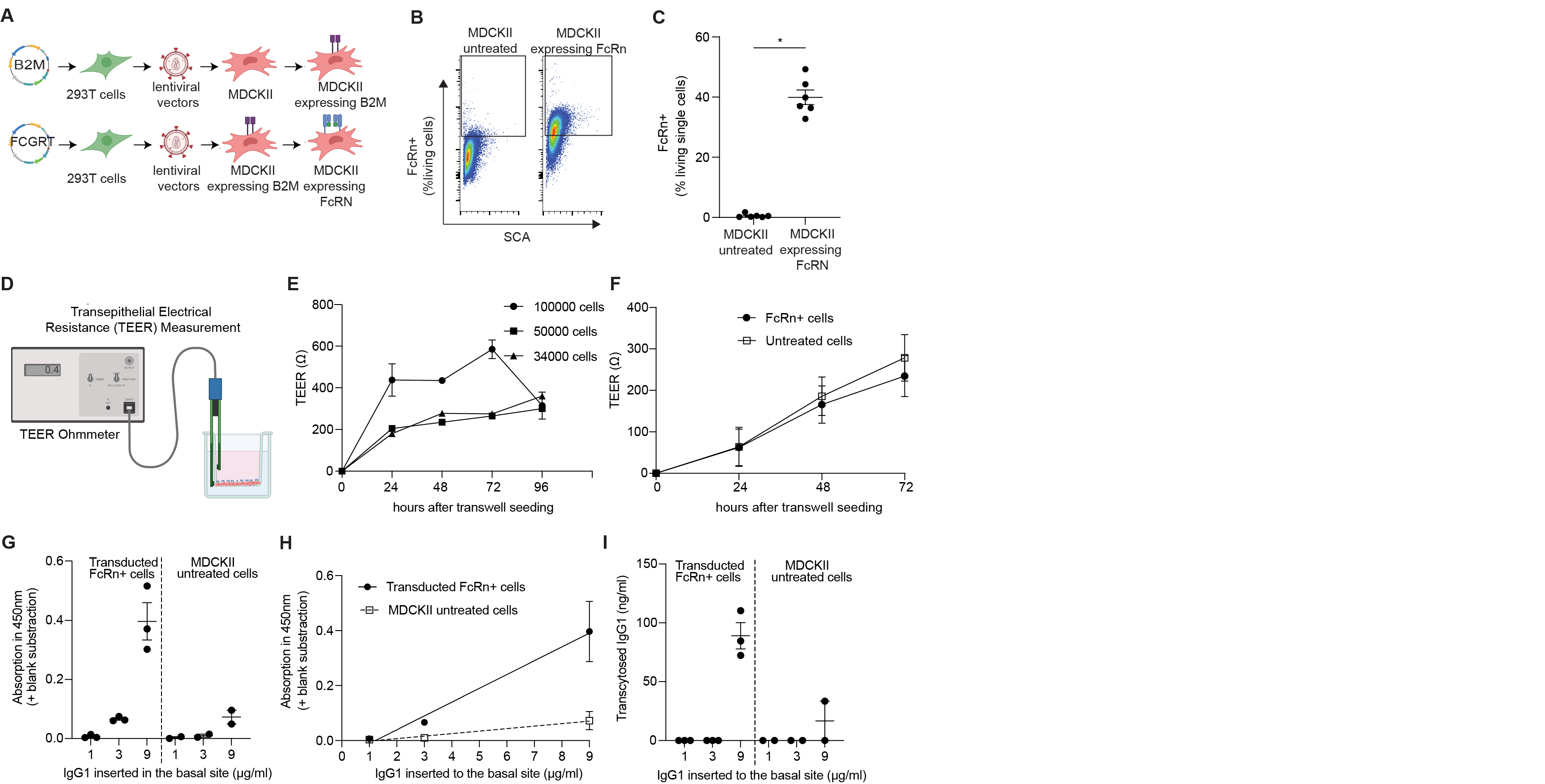

Fig. S11: Workflow for transduction and transcytosis assay setup with MDCKII cells. (A) Overview of the transduction process used to create MDCKII FcRn+ cells. (Figures crafted with Biorender). B2M; Beta-2 microglobulin, FCGRT; Fcγ receptor and transporter. (B) Flow cytometry plots illustrating untreated MDCKII cells versus transducted MDCKII FcRn+ cells, identified with an APC-conjugated anti-alpha-tag nanobody. (C) Percentage of FcRn-positive (FcRn+) cells within the viable population, determined via flow cytometry. Data from six independent cultures of untreated MDCKII cells and six cultures of transduced MDCKII FcRn+ cells. (D) Diagram of the setup for measuring transepithelial electrical resistance (TEER). (E) TEER measurements over time in 24-well Transwell plates seeded with different cell densities to find the optimal density for reaching a TEER plateau within 72 hours. (F) TEER over time in plates with either untreated MDCKII cells or FCRN+ cells at 50,000 cells per well, showing no significant differences. (G-I) Quantification of transcytosed IgG after 2 hours in the transwell system, comparing FCRN+ and untreated MDCKII cells with varying basal IgG concentrations. The 6μg/ml concentration was deemed optimal based on data shown in (H).

| Participants tested for Ab titers (n=38) | | | |
| --- | --- | --- | --- |
|  | 1^st^ Pregnancy | 2^nd^ Pregnancy | p value |
| **Maternal parameters** | | | |
| Age at birth (years; mean ± SD) | 31.8 ± 3.2 | 35.3 ± 3.5 | <0.0001 |
| Body Mass Index (kg/m^2^; mean ± SD) | 23.8 ± 4 | 24.4 ± 4.8 | 0.07 |
| - Underweight (<18.5 kg/m^2^; %) | 0 | 2.8 | 0.56 |
| - Normal weight (18.5–24.9 kg/m^2^; %) | 68.4 | 69.4 |  |
| - Overweight  (>25 kg/m^2^; %) | 31.6 | 27.8 |  |
| Educational Level |  |  |  |
| - School attendance <10 years (%) | 24.3 | 17.7 | 0.005 |
| - School attendance > 10 years (%) | 67.6 | 41.2 |  |
| - University Degree (%) | 8.1 | 41.1 |  |
| Delivery Mode |  |  |  |
| - Vaginal birth (%) | 81.6 | 81.6 | >0.99 |
| - C section (%) | 18.4 | 18.4 |  |
| **Neonate’s parameters** | | | |
| Gestational weeks at birth (mean ± SD) | 39.3 ± 1.4 | 39.3 ± 0.9 | 0.74 |
| - Term (≥37; %) | 97.4 | 100 | >0.99 |
| - Preterm (<37%; %) | 2.6 | 0 |  |
| Sex |  |  |  |
| - Male (%) | 60.5 | 46 | 0.25 |
| - Female (%) | 39.5 | 54 |  |
| Birth weight (g; mean± SD) | 3469.3 ± 544 | 3560.5 ± 438.4 | 0.18 |
| Height at birth (cm; mean± SD) | 52 ± 2.81 | 52.4 ± 2.4 | 0.38 |

Table S1: Demographic details of PRINCE study participants with two consecutive pregnancies.

**Table S2: Antibody titers and seropositivity prevalence among PRINCE participants who had two consecutive pregnancies and were tested for antibody titers.** The comparison of antibody titers between the first and second pregnancies was conducted using either a paired t-test or a Wilcoxon test, depending on the data distribution.

| **IgG Antibodies** | **Maternal serum** | | | | |
| --- | --- | --- | --- | --- | --- |
|  | 1^st^ pregnancy | | 2^nd^ pregnancy | | 1^st^ vs. 2^nd^pregnancy  p value |
|  | Seropositivity | Abs concentration  (mean ± SD) | Seropositivity | Abs concentration  (mean ± SD) |  |
| Influenza (RU/ml) | 38/38 | 85.79 ± 49.54 | 38/38 | 84.39 ± 44.95 | 0.77 |
| Diphtheriae (IU/ml) | 35/36 | 0.45 ± 0.43 | 35/36 | 0.36 ± 0.3 | 0.04 |
| B. pertussis (IU/ml) | 27/31 | 15.4 ± 11.7 | 27/31 | 18.24 ± 16.9 | 0.46 |
| Tetanus (IU/ml) | 25/25 | 2.01 ± 1.09 | 25/25 | 1.53 ± 0.67 | 0.003 |
| Measles (IU/l) | 36/36 | 914.17 ± 1272.14 | 36/36 | 802.09 ± 1052.98 | 0.77 |
| RSV (RU/ml) | 23/24 | 56.5 ± 41.18 | 24/24 | 66.33 ± 57.48 | 0.77 |
| Mumps (RU/ml) | 35/35 | 39.75 ± 33.51 | 35/35 | 39.86 ± 28.13 | 0.9 |
| Rubella (IU/ml) | 37/37 | 68.6 ± 51.93 | 37/37 | 66.16 ± 44.67 | 0.61 |
|  | **Cord serum** | | | | |
|  | 1^st^ pregnancy | | 2^nd^ pregnancy | | 1^st^ vs. 2^nd^pregnancy  p value |
|  | Seropositivity | Abs concentration  (mean ± SD) | Seropositivity | Abs concentration  (mean ± SD) |  |
| Influenza (RU/ml) | 38/38 | 115.07 ± 50.78 | 38/38 | 104.74 ± 44.96 | 0.05 |
| Diphtheriae (IU/ml) | 35/36 | 0.61 ± 0.6 | 35/36 | 0.45 ± 0.35 | 0.03 |
| B. perussis (IU/ml) | 27/31 | 23.61 ± 19.7 | 27/31 | 26.48 ± 26.11 | 0.51 |
| Tetanus (IU/ml) | 25/25 | 2.67 ± 1.53 | 25/25 | 2.18 ± 1.21 | 0.05 |
| Measles (IU/l) | 36/36 | 1490.35 ± 2174.9 | 36/36 | 1246.32 ± 1638.49 | 0.35 |
| RSV (RU/ml) | 23/24 | 98.66 ± 61.39 | 24/24 | 102.39 ± 70.84 | 0.69 |
| Mumps (RU/ml) | 35/35 | 59.33 ± 48.61 | 35/35 | 57.63 ± 44.22 | 0.63 |
| Rubella (IU/ml) | 37/37 | 109.44 ± 99.6 | 37/37 | 96. 24 ± 78.5 | 0.44 |

| **Exon** | **Appearing pseudo-SNP according to *IGHG1* reference** | **Type** | **DNA** | **Eu numbering changes - protein** | **Protein** | **Frequency**  **in study-**  **population**  **(%)** | **Reference**  **Frequency in non-Finnish**  **Europeans (%)** | **Reference**  **Frequency globally (%)** | **Beta**  **effect** | **Std.err** | **p value** |
| --- | --- | --- | --- | --- | --- | --- | --- | --- | --- | --- | --- |
| **Exon 1** | **rs17850094** | **Synonym** | **c.36A>G** | **Ala129=** | **Ala12=** | **29,9** | **0,4204** | **0,3094** | **-25,8** | **11,4** | **0,026** |
| **Exon 1** | **rs587637831** | **Missense** | **c.41C>G** | **p.Ser131Cys** | **p.Ser14Cys** | **37,3** | **0,147** | **0,1158** | **-22,7** | **10,7** | **0,038** |
| Exon 1 | rs745567365 | Missense | c.47A>G | p.Lys133Arg | p.Lys16Arg | 4,5 | 0,03103 | 0,02819 | -33,2 | 25,3 | 0,195 |
| Exon 1 | rs376989434 | Missense | c.58G>C | p.Gly137Arg | p.Gly20Arg | 22,4 | 0,0125 | 0,009422 | -21,2 | 12,80 | 0,103 |
| Exon 1 | rs372366008 | Missense | c.59G>A | p.Gly20Glu | p.Gly20Glu | 34,3 | 0,009825 | 0,008392 | -16,7 | 11,1 | 0,138 |
| Exon 1 | rs368400796 | Missense | c.61G>A | p.Gly138Ser | p.Gly21Ser | 34,3 | 0,01107 | 0,008929 | -16,7 | 11,1 | 0,138 |
| Exon 1 | rs587645157 | Synonym | c.234C>G | p.Thr195= | p.Thr78= | 7,5 | 0,00122 | 0,001306 | -15,4 | 22,3 | 0,494 |
| Exon 1 | rs587771912 | Missense | c.235C>A | p.Gln196Lys | p.Gln79Lys | 70,2 | 0,0009741 | 0,001174 | 1,8 | 11,6 | 0,878 |
| Exon 1 | rs587729878 | Missense | c.245T>C | p.Ile199Thr | p.Ile82Thr | 25,4 | 0,009618 | 0,01691 | -22,5 | 12,2 | 0,068 |
| Exon 1 | rs183696896 | Synonym | c.252C>T | p.Asn201= | p.Asn84= | 25,4 | 0,002875 | 0,02212 | -22,5 | 12,2 | 0,068 |
| **Exon 1** | **rs587772470** | **Synonym** | **c.255G>A** | **p.Val202=** | **p.Val85=** | **26,9** | **0,000479** | **0,001157** | **-24,5** | **11,8** | **0,042** |
| Exon 1 | rs1071803 | Missense | c.290A>G | p.Lys144Arg | p.Lys97Arg | 47,8 | 69,65 | 58,28 | 14,6 | 10,5 | 0,171 |
| Exon 3 | rs1042895 | Missense | c.536A>T | p.Tyr296Phe | p.Tyr179Phe | 4,5 | 0,04212 | 0,05745 | - | - | - |
| Exon 3 | rs1043249 | Synonym | c.594T>C | p.Asn315= | p.Asn198= | 14,9 | 0,3111 | 0,5821 | -22,1 | 15,3 | 0,154 |
| Exon 3 | rs369850034 | Missense | c.629C>G | p.Ala327Gly | p.Ala210Gly | 19,4 | 0,03111 | 0,02576 | -19,6 | 13,6 | 0,155 |
| Exon 3 | rs1043871 | Missense | c.637G>T | p.Ala330Ser | p.Ala213Ser | 19,4 | 0,005982 | 0,003717 | -19,6 | 13,6 | 0,155 |
| Exon 3 | rs1062730 | Missense | c.640C>T | pPro331Ser | pPro214Ser | 22,4 | 0,005982 | 0,00346 | -8,4 | 13 | 0,522 |
| Exon 4 | rs1050573 | Synonym | c.912C>T | p.Asn421= | p.Asn304= | 16,4 | 0,02303 | 0,0185 | -24,7 | 16,1 | 0,130 |
| Exon 4 | rs755034986 | Missense | c.713G>A | p.Arg355Gln | p.Arg238Gln | 20,9 | 0,01322 | 0,03688 | -17,6 | 13,3 | 0,188 |
| Exon 4 | rs1045853 | Missense | c.717T>G | p.Asp356Glu | p.Asp239Glu | 46,3 | 69,93 | 55,67 | 14,2 | 10,6 | 0,183 |
| Exon 4 | rs11621259 | Missense | c.721C>A | p.Leu358Met | p.Leu241Met | 44,8 | 69,8 | 55,36 | 14,3 | 10,6 | 0,182 |
| Exon 4 | rs58566624 | Synonym | c.768T>C | p.Tyr373= | p.Tyr256= | 28,4 | 0,5721 | 0,4843 | -12,6 | 11,9 | 0,292 |
| Exon 4 | rs17841087 | Synonym | c.870C>T | p.Tyr407= | p.Tyr290= | 58,2 | 55,47 | 45,99 | 14,1 | 10,7 | 0,191 |
| Intron 4 | rs1487627615 | intronic | c.985+12C>A | - | - | 3 | 0,001231 | 0,001054 | - | - | - |
| Intron 4 | rs12880412 | intronic | c.985+13C>G | - | - | 25,4 | 49 | 49,19 | -18,6 | 12,3 | 0,133 |
| **Exon 4** | **rs1063407** | **Missense** | **c.875A>G** | **p.Lys409Arg** | **p.Lys292Arg** | **26,9** | **0,004314** | **0,003474** | **-27,2** | **11,7** | **0,023** |
| Exon 4 | rs1050170 | Synonym | c.879C>A | p.Leu410= | p.Leu293= | 14,9 | 0,005511 | 0,003602 | -23,6 | 15,3 | 0,128 |
| Exon 4 | rs777509204 | Missense | c.904C>G | p.Gln419Glu | p.Gln302Glu | 1,5 | 0,06176 | 0,05326 | - | - | - |
| Exon 4 | rs370656573 | Synonym | c.684A>G | p.Glu345= | p.Glu228= | 13,4 | 0,005033 | 0,00772 | -21,4 | 14,7 | 0,149 |
| Exon 4 | rs113804727 | Missense | c.941C>G | p.Ala431Gly | p.Ala314Gly | 11,9 | 9,277 | 9,107 | -5,2 | 16,4 | 0,749 |
| Exon 4 | rs12879979 | Synonym | c.981T>C | p.Ser444= | p.Ser327= | 56,7 | 43,25 | 34,46 | 11,1 | 10,7 | 0,304 |
| Exon 4 | rs756753255 | Missense | c.983C>T | p.Pro445Leu | p.Pro328Leu | 13,43 | 0,03488 | 0,01624 | -24,7 | 16,1 | 0,130 |

Table S3: Pseudo-SNPs identified through Sanger sequencing, accompanied by regression analysis results. A linear regression using an additive genetic model was conducted. The beta effect, standard error, and p-value for each analysis are shown in the right columns.

| **Gene** | **Exon 1** | **Intron 1** | **Exon 2** | **Intron 2** | **Exon 3** | **Intron 3** | **Exon 4** |
| --- | --- | --- | --- | --- | --- | --- | --- |
| **IGHG1** | 95,25% | 93,61% | 55,56% | 97,46% | 95,15% | 93,81% | 96,83% |
| **IGHG2** | 97,63% | 95,41% | 53,49% | 95,76% | 93,64% | 94,85% | 96,83% |
| **IGHG3** | 96,27 | 94,64% | multiple hinge exons – exon intron structure different | | | | |
| **IGHG4** | 98,98% | 99,49% | 79,07% | 100% | 99,39% | 100% | 99,68% |

**Table S4: Representative analysis of a duplication with hybrid Exon 2.** *IGHG1* (ENST00000631539.1), *IGHG2* (ENST00000641095.1_11), *IGHG3* (ENST00000390551.6), *IGHG4* (ENST00000641978.1). For detailed Exon 2 structure, see also Fig.S6C.

| **IGHG1 copy** | **Individual 1** | **Individual 2** | **Individual 3** | **Individual 4** |
| --- | --- | --- | --- | --- |
| **Individual 1** |  | 100% | 100% | 100% |
| **Individual 2** | 100% |  | 100% | 100% |
| **Individual 3** | 100% | 100% |  | 100% |
| **Individual 4** | 100% | 100% | 100% |  |

**Table S5: Homology between the canonical IGHG1 copies across different individuals, based on long sequencing data.**

| **duplication copy** | **1.1** | **2.1** | **2.2** | **3.1** | **3.2** | **4.1** |
| --- | --- | --- | --- | --- | --- | --- |
| **1.1** |  | 99,3% | 99,4% | 99,89% | 99,4% | 99,4% |
| **2.1** | 99,3% |  | 99,13% | 99,4% | 99,13% | 99,13% |
| **2.2** | 99,4% | 99,13% |  | 99,29% | 100% | 100% |
| **3.1** | 99,89% | 99,4% | 99,29% |  | 99,29% | 99,29% |
| **3.2** | 99,4% | 99,13% | 100% | 99,29% |  | 100% |
| **4.1** | 99,4% | 99,13% | 100% | 99,29% | 100% |  |

**Table S6: Homology across the hybrid IGHG1/4 duplication and triplication copies in different individuals.**

|  | Non-carriers (n=55) | | Carriers (n=11) | p value |
| --- | --- | --- | --- | --- |
| Maternal age at delivery (years; mean ± SD) | 32.4±2.9 | | 33.5 ± 4.1 | 0.31 |
| Maternal Body Mass Index (kg/m^2^; mean ± SD) | 24.4±4 | | 23.3± 2.6 | 0.36 |
| - Underweight (<18.5 kg/m^2^; %) | 0 | | 0 | 0.12 |
| - Normal weight (18.5–24.9 kg/m^2^; %) | 79.7 | | 54.5 |  |
| - Overweight (>25 kg/m^2^; %) | 20.3 | | 45.5 |  |
| Delivery mode |  | |  |  |
| - Vaginal birth (%) | 83.3 | | 68.8 | 0.28 |
| - C section (%) | 16.7 | | 31.3 |  |
| Neonatal gestational age at birth (mean ± SD) | 39.6 ± 1.3 | | 38.9 ± 1.4 | 0.15 |
| - Term (≥ 37^th^ week %) | 100 | | 100 | 0.99 |
| - Preterm (< 37%; %) | 0 | | 0 |  |
| Fetal sex |  | |  |  |
| - Male (%) | 48.1 | | 45.5 | 0.99 |
| - Female (%) | 51.9 | | 54.5 |  |
| Neonatal birth weight (g; mean ± SD) | 3465±508.6 | | 3416.7±469 | 0.77 |
| Neonatal height at birth (cm; mean ± SD) | 51.9 ± 2.6 | | 50.9 ± 2.3 | 0.27 |
| Participants with pregnancy complications (%) | 27.8 | | 35.3 | 0.55 |
| - Gestational hypertension (%) | 1.85 | | 0 |  |
| - Preeclampsia (%) | 0 | | 0 |  |
| - HELLP-syndrome (%) | 1.85 | | 0 |  |
| - Gestational diabetes (%) | 0 | | 9.1 |  |
| - Miscarriage (%) | 0 | | 0 |  |
| - Premature contractions (%) | 3.7 | | 4.8 |  |
| - Preterm labor (%) | 0 | | 0 |  |
| - Infection (%) | 9.25 | | 27.3 |  |
| - Other (%) | 16.6 | | 18.2 |  |
| Participants with chronic diseases | 20.8 | | 27.3 | 0.69 |
| - Hypothyroidism | 9.3 | | 0 |  |
| - Asthma | 7.4 | | 9.1 |  |
| - Other | 3.7 | | 18.2 |  |

Table S7: Demographic characteristics of pregnant women with chimeric IgG1/IgG4 alleles (carriers) compared to those with wild-type alleles (non-carriers).

| Antibody ID | Heavy / Light chain | Amino acid sequence |
| --- | --- | --- |
| antibody carrying canonical *IGHG1* sequence | Heavy chain | EVQLVESGGGLVQPGGSLRLSCAASGFSFSSYWMTWVRQAPGKGLEWVANIKQYGSEKYY  VDSVKGRFTISRDNAKNSLYLQMNSLRDDDTAVYYCARMGSYLDTYYYHYGMDVWGQGTT  VTVSASTKGPSVFPLAPSSKSTSGGTAALGCLVKDYFPEPVTVSWNSGALTSGVHTFPAVLQSS  GLYSLSSVVTVPSSSLGTQTYICNVNHKPSNTKVDKKVEPKSCDKTHTCPPCPAPELLGG  PSVFLFPPKPKDTLMISRTPEVTCVVVDVSHEDPEVKFNWYVDGVEVHNAKTKPREEQYN  STYRVVSVLTVLHQDWLNGKEYKCKVSNKALPAPIEKTISKAKGQPREPQVYTLPPSRDE  LTKNQVSLTCLVKGFYPSDIAVEWESNGQPENNYKTTPPVLDSDGSFFLYSKLTVDKSRW  QQGNVFSCSVMHEALHNHYTQKSLSLSPELQLEESCAEAQDGELDGLWTTITIFITLFLL  SVCYSATVTFFKVKWIFSSVVDLKQTIIPDYRNMIGQGA |
| antibody carrying marker pseudo-SNPs | Heavy chain | EVQLVESGGGLVQPGGSLRLSCAASGFSFSSYWMTWVRQAPGKGLEWVANIKQYGSEKYY  VDSVKGRFTISRDNAKNSLYLQMNSLRDDDTAVYYCARMGSYLDTYYYHYGMDVWGQGTT  VTVSASTKGPSVFPLAPCSKSTSGGTAALGCLVKDYFPEPVTVSWNSGALTSGVHTFPAVLQSS  GLYSLSSVVTVPSSSLGTQTYICNVNHKPSNTKVDKKVEPKSCDKTHTCPPCPAPELLGG  PSVFLFPPKPKDTLMISRTPEVTCVVVDVSHEDPEVKFNWYVDGVEVHNAKTKPREEQYN  STYRVVSVLTVLHQDWLNGKEYKCKVSNKALPAPIEKTISKAKGQPREPQVYTLPPSRDE  LTKNQVSLTCLVKGFYPSDIAVEWESNGQPENNYKTTPPVLDSDGSFFLYSRLTVDKSRW  QQGNVFSCSVMHEALHNHYTQKSLSLSPELQLEESCAEAQDGELDGLWTTITIFITLFLL  SVCYSATVTFFKVKWIFSSVVDLKQTIIPDYRNMIGQGA |
| antibody carrying all detected pseudo-SNPs | Heavy chain | EVQLVESGGGLVQPGGSLRLSCAASGFSFSSYWMTWVRQAPGKGLEWVANIKQYGSEKYY  VDSVKGRFTISRDNAKNSLYLQMNSLRDDDTAVYYCARMGSYLDTYYYHYGMDVWGQGTT  VTVSASTKGPSVFPLAPCSRSTSRSTAALGRLVKDYFPEPVTVSWNSGALTSGVHTFPAVLQSS  GLYSLSSVVTVPSSSLGTKTYTCNVNHKPSNTKVDKRVEPKSCDKTHTCPPCPAPELLGG  PSVFLFPPKPKDTLMISRTPEVTCVVVDVSHEDPEVKFNWYVDGVEVHNAKTKPREEQFN  STYRVVSVLTVLHQDWLNGKEYKCKVSNKGLPSSIEKTISKAKGQPREPQVYTLPPSQEE  MTKNQVSLTCLVKGFYPSDIAVEWESNGQPENNYKTTPPVLDSDGSFFLYSRLTVDKSRW  QEGNVFSCSVMHEGLHNHYTQKSLSLSLELQLEESCAEAQDGELDGLWTTITIFITLFLL  SVCYSATVTFFKVKWIFSSVVDLKQTIIPDYRNMIGQGA |
| All three variants | Light chain | MAWMMLLLTLLAHCTGSWAQSVLTQPPSLSGAPGQRVTISCTGSSSNIGADYDVYWYQHL  PGTAPKLLMYGDGYRPSGVPDRFSGSKSGTSASLAITGLQAEDEADYYCQSYDSSLSRRV  VFGGGTKLTVLGQPKAAPSVTLFPPSSEELQANKATLMCLISDFYPGAVTVAWKADSSPV  KAGVETTTPSKQSNNKYAASSYLSLTPEQWKSHRSYSCQVTHEGSTVEKTVAPTECS |

Table S3: Pseudo-SNPs identified through Sanger sequencing, accompanied by regression analysis results. A linear regression using an additive genetic model was conducted. The beta effect, standard error, and p-value for each analysis are shown in the right columns.
